# Single-cell profiling reveals immune aberrations in progressive idiopathic pulmonary fibrosis

**DOI:** 10.1101/2023.04.29.23289296

**Authors:** Avraham Unterman, Amy Y. Zhao, Nir Neumark, Jonas C. Schupp, Farida Ahangari, Carlos Cosme, Prapti Sharma, Jasper Flint, Yan Stein, Changwan Ryu, Genta Ishikawa, Tomokazu S. Sumida, Jose L. Gomez, Jose Herazo-Maya, Charles S. Dela Cruz, Erica L. Herzog, Naftali Kaminski

## Abstract

**Rationale:** Changes in peripheral blood cell populations have been observed but not detailed at single-cell resolution in idiopathic pulmonary fibrosis (IPF).

**Objectives:** To provide an atlas of the changes in the peripheral immune system in stable and progressive IPF.

**Methods:** Peripheral blood mononuclear cells (PBMCs) from IPF patients and controls were profiled using 10x Chromium 5’ single-cell RNA sequencing (scRNA-seq). Flow cytometry was used for validation. Protein concentrations of Regulatory T-cells (Tregs) and Monocytes chemoattractants were measured in plasma and lung homogenates from patients and controls.

**Measurements and Main Results:** Thirty-eight PBMC samples from 25 patients with IPF and 13 matched controls yielded 149,564 cells that segregated into 23 subpopulations, corresponding to all expected peripheral blood cell populations. Classical monocytes were increased in progressive and stable IPF compared to controls (32.1%, 25.2%, 17.9%, respectively, p<0.05). Total lymphocytes were decreased in IPF vs controls, and in progressive vs stable IPF (52.6% vs 62.6%, p=0.035). Tregs were increased in progressive IPF (1.8% vs 1.1%, p=0.007), and were associated with decreased survival (P=0.009 in Kaplan-Meier analysis). Flow cytometry analysis confirmed this finding in an independent cohort of IPF patients. Tregs were also increased in two cohorts of lung scRNA-seq. CCL22 and CCL18, ligands for CCR4 and CCR8 Treg chemotaxis receptors, were increased in IPF.

**Conclusions:** The single-cell atlas of the peripheral immune system in IPF, reveals an outcome-predictive increase in classical monocytes and Tregs, as well as evidence for a lung-blood immune recruitment axis involving CCL7 (for classical monocytes) and CCL18/CCL22 (for Tregs).

## Introduction

The role of the immune system in idiopathic pulmonary fibrosis (IPF), a chronic interstitial lung disease that leads to progressive lung scarring and death (1), is not fully resolved (2). Nearly every immune cell type has been suggested to play a role in the disease, including monocytes, macrophages, T-cells and B-cells (2–4), but immunosuppressive treatments increase mortality in IPF patients (5). We have previously identified and validated a 52-gene expression signature in peripheral blood mononuclear cells (PBMCs) bulk RNA samples, predictive of outcome in patients with IPF (6, 7), a finding that has led to the discovery that increased monocytes are associated with mortality in IPF (8, 9). However, a detailed profile of peripheral immune cell composition and aberrations in IPF is still missing.

Recent advancements in single-cell RNA sequencing technology (scRNA-seq) provide an exciting opportunity to study the gene expression profiles of individual cells (10). Their application to tissues obtained from patients with end stage pulmonary fibrosis led to substantial insights, and highlighted changes in immune cell populations in the lung (11, 12), but a detailed catalogue of these changes in the peripheral blood is still missing. Here we provide an atlas of the peripheral immune system in IPF according to disease presence and progression. ScRNA-seq analysis of PBMCs obtained from stable and progressive IPF patients, as well as matched control subjects reveals that classical and intermediate monocytes are increased in progressive IPF, whereas total lymphocytes are decreased. The opposite is true for regulatory T cells (Tregs) which are activated and increased in progressive IPF, both in the peripheral blood and in two cohorts of lung scRNA-seq. We demonstrate that chemoattractants potentially involved in monocytes and Tregs recruitment are increased in plasma and lung tissue homogenates of IPF patients, suggesting a lung-blood immune recruitment model in IPF. Results can be further explored through our user-friendly data-mining website (http://ildimmunecellatlas.com).

## Methods

### Settings, patients, and samples

The study was performed on deidentified, cryopreserved PBMC samples of IPF patients and matched controls, previously obtained with informed consent on protocol approved by the Institutional Review Board at the Yale School of Medicine (#1307012431).

IPF subjects were enrolled between 2012 and 2015 using an IPF diagnosis based on consensus guidelines that were current at the time of enrollment (13) rendered through expert multidisciplinary discussion. Demographic and clinical data were obtained at the time of the blood draw. IPF patients were all untreated at the time of the blood draw. They were followed for 36 months for time-to-event analysis, where death from any cause was considered an event. Those subjects alive at the end of the 36-month period were considered “Stable”, while those who died were termed “Progressive”. Figure 1A shows the Kaplan-Meyer curves for both groups of IPF patients. A group of demographically matched healthy controls without known fibrosing, inflammatory, or other lung disease was studied as well. Stable and Progressive IPF patients were matched for age, sex, race, smoking status, and lung function, while IPF and control subjects were matched for age, sex, race, and smoking status (Table 1). A sample code was given to each sample, containing a letter (“S” for stable IPF, “P” for progressive IPF, and “C” for control subjects) and a number between 01 and 40. Two samples, one control (C28) and one progressive IPF (P13), were excluded following QC due to low sample quality. The final cohort for analysis consisted of 38 samples: 13 stable IPF, 12 progressive IPF and 13 controls (Figure 1).

**Figure 1:**
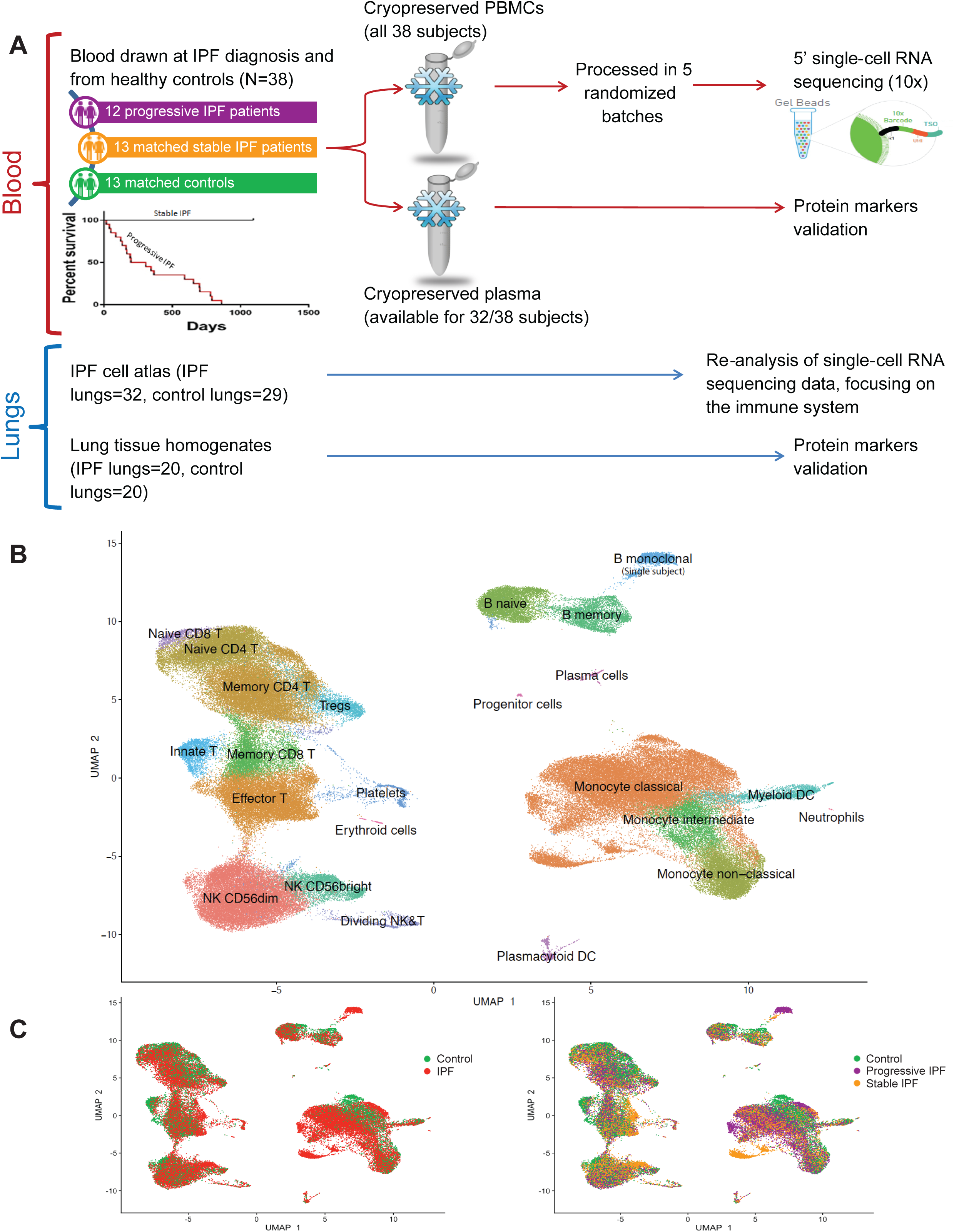
Study design and single-cell clustering results. A. Banked, cryopreserved PBMCs from 12 progressive IPF patients, 13 matched stable IPF patients and 13 matched control subjects were included in this study. Blood had been drawn at the time of IPF diagnosis and was processed into cryopreserved PBMCs and plasma. IPF patients were followed for 36 months after the blood draw. Those subjects alive at the end of the 36 months period were considered “Stable”, while those who died were termed “Progressive”, as depicted in the Kaplan-Meyer curve. Cryopreserved PBMCs were processed in 5 randomized batches and subjected to 5’ scRNA-seq while plasma was used to determine the level of relevant cytokines and chemokines. Re-analysis of lung scRNA-seq data and cytokine levels in lung tissue homogenates supplemented the blood-based data. B. UMAP representation of 149,564 cells parceled into 23 cell types. All expected cell types were identified. C. Same UMAP as in panel B, with cells color coded according to disease and disease progression. Abbreviations: PBMCs peripheral blood mononuclear cells, DC dendritic cells, NK natural killer cells.

**Table 1:** Baseline characteristics of IPF patients and control subjects. Abbreviations: IPF idiopathic pulmonary fibrosis, SD standard deviation, FVC forced vital capacity, DLCO diffusion capacity of the lungs to carbon monoxide, NA not available.

|  | Progressive IPF | Stable IPF | P value | All IPF | Controls | P value |
| --- | --- | --- | --- | --- | --- | --- |
| N | 12 | 13 |  | 25 | 13 |  |
| Age mean (SD), years | 70.8 (7.0) | 69.9 (6.1) | 0.76 | 70.3 (6.4) | 71.0 (4.2) | 0.70 |
| Male sex | 83% | 77% | 0.69 | 80% | 77% | 0.83 |
| Caucasian race | 92% | 100% | 0.29 | 96% | 100% | 0.46 |
| Current/former smoker | 67% | 77% | 0.57 | 72% | 69% | 0.86 |
| FVC, % predicted | 69.83 | 76.85 | 0.20 | 73.48 | NA | NA |
| DLCO, % predicted | 37.67 | 44.31 | 0.20 | 41.12 | NA | NA |

### PBMC isolation and cryopreservation

We used previously obtained, deidentified, cryopreserved PBMCs that had been isolated from whole blood using density gradient centrifugation, according to the following protocol. Blood was centrifuged at 400g for 8 minutes at room temperature to separate cells from plasma. The plasma was subsequently aliquoted and stored at −80°C, and the remaining sample was diluted 1:2 with sterile PBS. Next, 12 ml layer of Histopaque 1077 (Sigma Aldrich) was overlaid with the diluted sample, and the mixture was subsequently centrifuged at 1200g for 30 minutes at 12°C. The buffy coat was then transferred to a new tube, diluted with PBS up to 50 ml, gently mixed and centrifuged at 450g for 10 minutes at 4°C. The cells were then washed two times with 50 ml PBS and one time in 10 ml PBS, with each wash followed by 450g centrifugation for 10 minutes at 4°C. For cryopreservation, at least 2*10^6^ cells per aliquot were resuspended in 70% DMEM, 20% FBS and 10% DMSO as the cryopreservation solution. Cryovials were placed in a freezing container (Mr. Frosty) and transferred immediately to −80°C for at least 24 hours, before transferring to liquid nitrogen for long-term storage.

### Sample preparation and single-cell barcoding

To minimize batch effects, samples were processed in five randomized and balanced batches of 8, each containing 2-3 samples of each of the 3 patient categories: stable IPF, progressive IPF, and controls (Table E1 in the online supplement). Samples were thawed in a water bath at 37°C for ∼2 min without agitation and removed from the water bath when a tiny ice crystal still remains. After thawing, cells were gently transferred to a 50 mL conical tube using a wide-bore pipette tip, the cryovial was rinsed with cold growth medium (10% FBS in DMEM) to recover leftover cells, and the rinse medium was added dropwise (1 drop per 5 sec) to the 50 mL conical tube while gently shaking the tube. Next, we conducted 5 serial dilutions with cold growth medium slowly added at 1:1 ratio, achieving a final volume of 32 mL. The cells were then centrifuged at 300 x g for 5 minutes at 4°C, and the supernatant was removed without disrupting the cell pellet. The pellet was resuspended in 1X PBS with 0.04% BSA, and the sample was filtered with a 40 μM strainer. Cell concentration was determined using Trypan blue staining with a Countess automated cell counter (ThermoFisher). One million cells from each sample in batch 1 were set aside for CyTOF validation (as further described below). Cells from each sample were loaded onto the 10x Chromium Chip A, according to the manufacturer’s user guide (Chromium Single Cell V(D)J Reagent Kits User Guide, document number CG000086, revision J, November 2018), aiming for recovery of 10,000 cells per lane.

### 5’ cDNA libraries preparation and sequencing

The loaded Chip A was placed in the 10x Chromium controller to create Gel Beads-in-emulsion (GEMs). The next steps were carried out according to the manufacturer’s user guide, including GEM-RT incubation, post GEM-RT Dynabead cleanup, and cDNA amplification. 5’ cDNA libraries were then sequenced on an Illumina Hiseq 4000 platform. Raw sequencing data will be available in GEO upon peer-reviewed publication.

### Data processing of raw sequencing reads

Raw sequencing reads were demultiplexed using CellRanger mkfastq pipeline to create FASTQ files. Next, CellRanger count pipeline (v3.0.2) was employed in order to perform alignment (using STAR), filtering, barcode counting, and UMI counting. We have used GRCh38 as the genome reference. We then used the CellRanger aggr pipeline (v3.0.2) to aggregate the outputs of multiple samples generated by CellRanger count, so that all samples will have the same effective sequencing depth.

### Data analysis using Seurat package

Seurat package (14, 15) (v3.1) was used for all downstream analyses. 10x gene expression aggregated matrix was converted into a Seurat object. Genes expressed by less than 3 cells were filtered out. Cells with mitochondrial gene percentages higher than 12% and cells with less than 1000 Unique Molecular Identifiers (UMIs) were excluded from the study to filter out dead and dying cells. After these filtering steps, the gene-barcode matrix contained 23,462 genes and 153,162 barcoded cells.

Gene expressions were normalized using Seurat’s “LogNormalize” method. The “FindVariableFeatures” function selected the 3,000 genes with the highest variance to mean ratio using the “vst” method. Due to the relative paucity of single-subject effects we decided not to use anchoring and data integration. The data was then scaled with the “ScaleData” function, with regression according to mitochondrial gene percentage.

Principal Component Analysis (PCA) was performed, and the first 30 Principal Components (PCs) were used in the “FindNeighbors” algorithm. Unsupervised Louvain clustering algorithm in “FindClusters” function yielded 35 clusters at 0.8 resolution (Figure E1A in the online supplement) and was followed by manual annotation of clusters into cell types based upon specific cell markers (Figure E1B). Five doublet clusters (#19, 27, 28, 29, 31) were then removed, and the final Seurat object for analysis consisted of 149,564 cells across 30 clusters aggregated into 23 cell types (Figure 1B). Cell type annotation was validated against an automated annotation algorithm (SingleR package, Figure E2).

### Cell Type Proportions Analysis

For each subject, the number of cells within a given cell type was normalized by the subject’s total number of cells. For each cell type, cell proportions were plotted in a boxplot, with subjects divided into the following groups: controls, stable IPF patients, and progressive IPF patients. We used the two-tailed T test for cell proportion comparisons.

### Differential Gene Expression Analysis and Heatmap visualization

The “FindMarkers” function in Seurat was used to identify differentially expressed genes (DEGs) between IPF and controls and between stable and progressive IPF. Wilcoxon rank sum test was used. Genes were ranked by absolute log*_e_* (ln) fold-change (logFC), and those with p-values > 0.05 (adjusted for multiple comparisons) were removed.

DEGs were visualized as heatmaps which were generated by using the ComplexHeatmap package (16). Cell types were binned into monocytes, CD4^+^ T cells, CD8^+^ T cells, and B cells and “FindMarkers” distinguished DEGs for each cell type bin. Genes with greater than 0.3 absolute logFC were included in visualization and EnrichR pathway analysis, excluding specific T cell receptor and B cell receptor genes. Samples for the IPF versus Control were hierarchically clustered.

### HLA type II Gene List Score

Seurat Function “AddModuleScore” was used to combine the expression of genes to create the HLA type II score (*HLA-DRA, HLA-DQA1, HLA-DPA1, HLA-DRB1, HLA-DPB1, HLA-DRB5, HLA-DQB1, HLA-DMA, HLA-DMB*).

### Reanalysis of lung scRNA-seq datasets to determine Treg frequency

The immune cells from Habermann et al (17) were re-processed using Seurat. Cells with mitochondrial gene percentages higher than 25% and cells with less than 1000 unique genes (“features”) were excluded. Gene expressions were normalized using Seurat’s “LogNormalize” method. The data was then scaled with the “ScaleData” function, with regression according to mitochondrial gene percentage. PCA was performed, and the first 40 PCs were used in the “FindNeighbors” algorithm. Unsupervised Louvain clustering algorithm in “FindClusters” function was employed with 0.8 resolution and was followed by manual annotation to detect all clusters of T cells, including Tregs. A boxplot of Tregs as percent of all T cells was created based on this analysis.

For Tregs frequency analysis in Adams et al (18), we used their original annotation of Tregs and T cells. A boxplot of Tregs as percent of all T cells was created based on these data.

### Cytometry by Time-Of-Flight (CyTOF)

We used mass cytometry (CyTOF) to validate the scRNA-seq based cell counts in some of the PBMC samples. One million cells per sample were used. Cells were incubated for 45 minutes on ice with a cocktail of metal-conjugated antibodies (Table E4). Next, the cells were washed and subsequently treated with 10 µM Cisplatin (Sigma-Aldrich, St. Louis, MO) for 5 minutes at room temperature. The cells were next washed and subsequently treated with Cell-ID 125 nM Intercalator-Ir (Fluidigm), fixed and permeabilized using Foxp3 Staining Buffer Set overnight at 4°C. Following the overnight incubation, the cells were washed and centrifuged at 800g for 10 minutes at 4°C. Immediately prior to data acquisition, 2.5-5*10^5^ cells were resuspended in Milli-Q water with 50% EQ beads (Fluidigm) and analyzed on CyTOF 2 machine (Fluidigm).

CyTOF-derived fcs files were processed using the bead-based Normalizer Release R2013a (19). Normalized files were then processed in Cytobank (https://premium.cytobank.org/) using gates to select singlets, remove beads and identify live cells. Events identified using this workflow were exported if there were more than 2000 live cells and processed further using the R package cytofkit version 1.12.0 (20). The Rphenograph function in cytofkit was implemented to cluster cells using cytofAsinh method, with the tsne dimensionality reduction method applied on 80000 events, using k=40. Files were merged using all events. CD45, CD235, CD19, CD45RA, CD8a, CD20, CD16, CD194/CCR4, CD123, IgD, CD4, CD183/CXCR3, CD3, CD11c, CD14, CD45RO, CD27, CD196/CCR6, CD25, CCR7/CD197, CD38, HLA-DR, CD56, and CD61 markers were used in this model. Resulting clusters were manually curated and merged after review of surface marker profiles.

### Flow cytometry

Flow cytometry validation was performed using an existing dataset comprised of PBMCs from 20 stable and 18 progressive IPF patients, different from the ones used for scRNA-seq. The original publication can be found here (21). We used a two-sided Mann-Whitney test to compare Treg proportions between progressive and stable IPF.

### Plasma samples protein analysis

Cryopreserved plasma samples from the same blood draw as the PBMC samples used for scRNA-seq, were available for 32 out of the 38 patients (9 controls, 12 stable IPF, and 11 progressive IPF). Plasma samples were thawed in a water bath and protein levels were measured using a custom-made U-PLEX Meso Scale Discovery (MSD) multiplex assay according to manufacturer’s instructions, using a MESO QuickPlex SQ 120 machine. The following proteins were measured using U-PLEX: M-CSF, G-CSF, GM-CSF, I-309 (CCL1), MCP-1 (CCL2), MCP-2 (CCL8), MCP-3 (CCL7), MCP-4 (CCL13), MDC (CCL22), and TARC (CCL17). Levels of PARC (CCL18) were measured using a Simple Plex kit on an Ella machine (ProteinSimple). Results were statistically analyzed on Prism 8, and P-values<0.05 were considered significant. We first conducted one-way ANOVA to determine significance across controls, stable and progressive IPF. Next, we applied the Tukey’s multiple comparisons test to determine if any of the three comparisons (control vs stable, stable vs progressive, control vs progressive) is statistically significant.

### Lung tissue homogenates protein analysis

Lung tissue homogenates were produced using a rotor-stator homogenizer from frozen IPF and control lung tissues, that were collected from explanted lungs and refuted donors, respectively. Protein levels were measured using a custom-made U-PLEX Meso-Scale Discovery (MSD) multiplex assay according to manufacturer’s instructions, using a MESO QuickPlex SQ 120 machine. The proteins measured using U-PLEX were the same as listed above for the plasma samples. Levels of PARC (CCL18) were measured using a Simple Plex kit on an Ella machine (ProteinSimple). Total protein concentration in each sample was measured with the Pierce BCA protein assay kit (ThermoFisher, Catalog No. 23225) in order to normalize the protein levels results. Unpaired T-test was used to compare IPF and control protein levels. Results were statistically analyzed on Prism 8, and P-values<0.05 were considered significant.

## Results

### PBMC subpopulations shift in association with disease presence and severity

We performed scRNA-seq of cryopreserved PBMCs obtained at the time of IPF diagnosis from 12 progressive IPF patients, 13 matched stable IPF patients, and 13 matched controls without known respiratory illness (Figure 1A). Progressive IPF patients differed from stable ones based on whether they died within 36 months of follow-up. Table 1 portrays the baseline characteristics of these three groups of subjects. The final cohort for analysis consisted of 149,564 cells across 30 clusters aggregated into 23 cell types (Figure 1B). All expected cell types were detected across all three patient groups (Figure 1C).

Alterations in cell type abundance were detected across control, stable, and progressive samples. There was a stepwise increase in mean counts of all monocytes (27.1% in controls, 33.5% in stable, and 43.3% in progressive IPF; Figure 2A), classical monocytes (17.9%, 25.2%, and 32.1%; Figure 2B), and intermediate monocytes (1.9%, 3.0%, and 4.7%). No difference in non-classical monocytes was noted between the three groups. Opposite to the increase in monocytes, combined levels of lymphocytes (T, B, and NK) exhibited a stepwise decrease (69.5%, 62.6%, and 52.6%; Figure 2A). Notable changes in the T lymphocytes compartment included a decrease in naïve CD4 and CD8 T cells, as well as effector T cells in progressive IPF (Figure 2B). In contrast, regulatory T cells (Tregs) were increased in progressive vs stable IPF (1.8% vs 1.1%, p=0.007). CyTOF-based cell counts were conducted as validation for a subset of our PBMC samples and were concordant with the scRNA-seq based cell counts (mean correlation coefficient 0.99; see Figure E3 in the online supplement).

**Figure 2:**
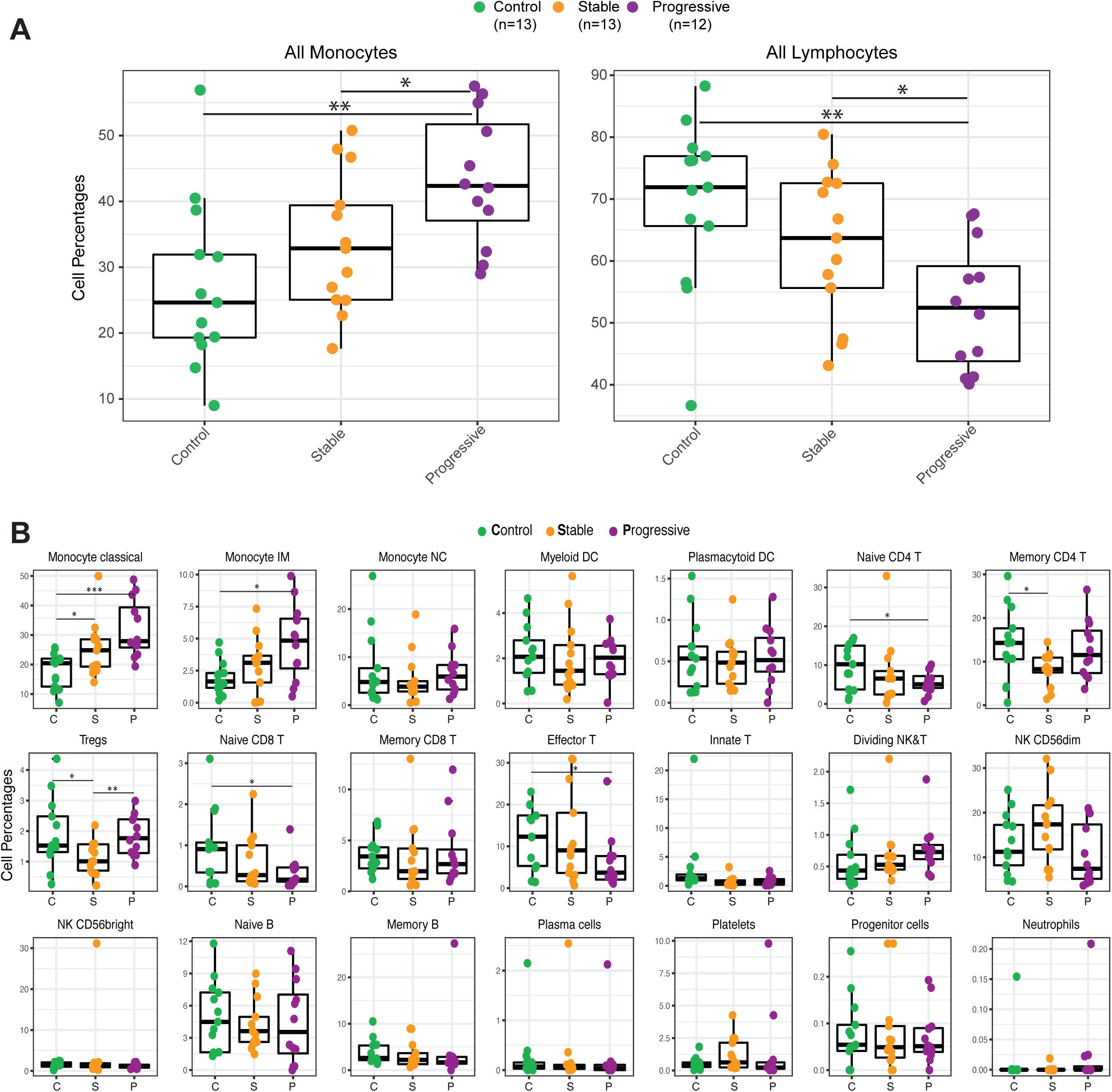
Shifts in PBMC subpopulations with disease severity. A. Boxplots showing scRNA-seq based cell proportions for all monocytes and all lymphocytes (as % of all PBMCs), grouped by control, stable, and progressive IPF. The results are depicted in boxplots, in which the value for each subject is represented by a dot, the upper and lower bounds represent the 75% and 25% percentiles, respectively. The center bars indicate the medians, and the whiskers denote values up to 1.5 interquartile ranges above the 75% or below the 25% percentiles. B. Similar boxplots showing scRNA-seq based cell proportions for each cell type. Abbreviations: DC dendritic cells, IM intermediate, NC non-classical, NK natural killer. * p<0.05, ** p<0.01.

We did not find a distinct population of COL1A1^+^CD45^+^ cells (22, 23). In our cohort COL1A1^+^CD45^+^ cells were rare (0.03% of cells), scattered across the T, B and monocyte compartments, not enriched in IPF patients, and did not form their own cluster (Figure E4).

### Differences in PBMC gene expression profile between IPF and controls

Gene expression shifts in IPF (stable and progressive patients combined, N=25) compared to controls (N=13) are depicted in Figure 3. An aberrant myeloid profile was evident in IPF relative to control subjects, with an increase of *S100A8* and *S100A12* and a decrease in HLA type II transcripts (Figure 3A-B; see Table E2 for a full list of genes and adjusted P values). This partly resembles the myeloid phenotype recently observed in progressive COVID-19 (24, 25).

**Figure 3:**
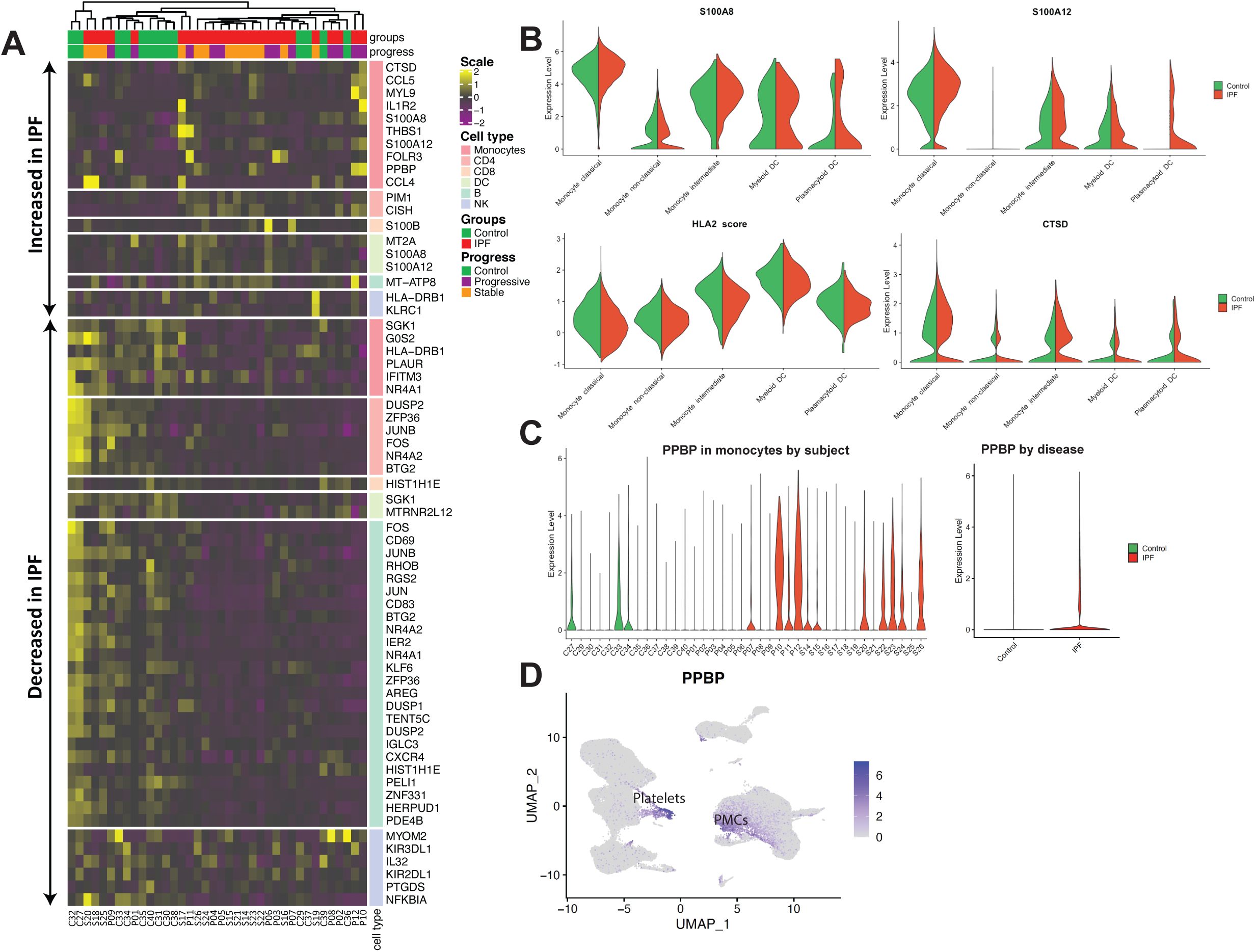
Differences in PBMC gene expression profile between IPF and controls. A. Heatmap showing the top differentially expressed genes (log2FC > 0.3, adjusted P values < 0.05) for each major cell type, comparing IPF patients to matched controls. The upper part of the heatmap depicts genes that are increased in IPF compared to control subjects (marked “increased in IPF”). Hierarchical clustering highlights the inter-subject variability in gene expression, leading to a mix of IPF and control subjects (color coded in upper panel). B. Violin plots focusing on selected genes that differ in monocytes and DCs between IPF and controls. *S100A8*, *S100A12* and *CTSD* are increased in IPF while a composite score of HLA class II transcripts is decreased. C. Violin plots depicting increased expression of the platelet-specific *PPBP* gene in monocyte clusters of IPF vs. controls. D. UMAP of *PPBP* expression in PBMCs, demonstrating increased expression in a subset of monocytes, potentially attributable to higher platelet-monocyte complexes (PMCs) formation in IPF. Abbreviations: DC dendritic cells, NK natural killer, PMCs platelet-monocyte complexes.

Expression of *PPBP*, a specific platelet gene, was increased in monocyte clusters of IPF vs. controls, although inter-subject variability existed (Figure 3C-D). Platelets, especially when activated, may adhere to monocytes, creating platelet-monocyte complexes (26). Two clusters of platelet-monocyte complexes were identified in our data, and were enriched in IPF (P=0.03; Figure E5), possibly reflecting an increase in platelet activation in IPF (27) with enhanced adherence to monocytes. Furthermore, levels of *PPBP* were also increased in pulmonary fibrosis vs control monocytes and macrophages in two datasets of lung scRNA-seq (17, 18) (Figure E5).

### Gene expression changes differentiate PBMCs of stable from progressive patients

Subpopulations of PBMCs from stable patients differed in gene expression from those of progressive patients (Figure 4A, Table E3). We conducted a gene enrichment pathway analysis to better characterize those differences (Figure E6).

**Figure 4:**
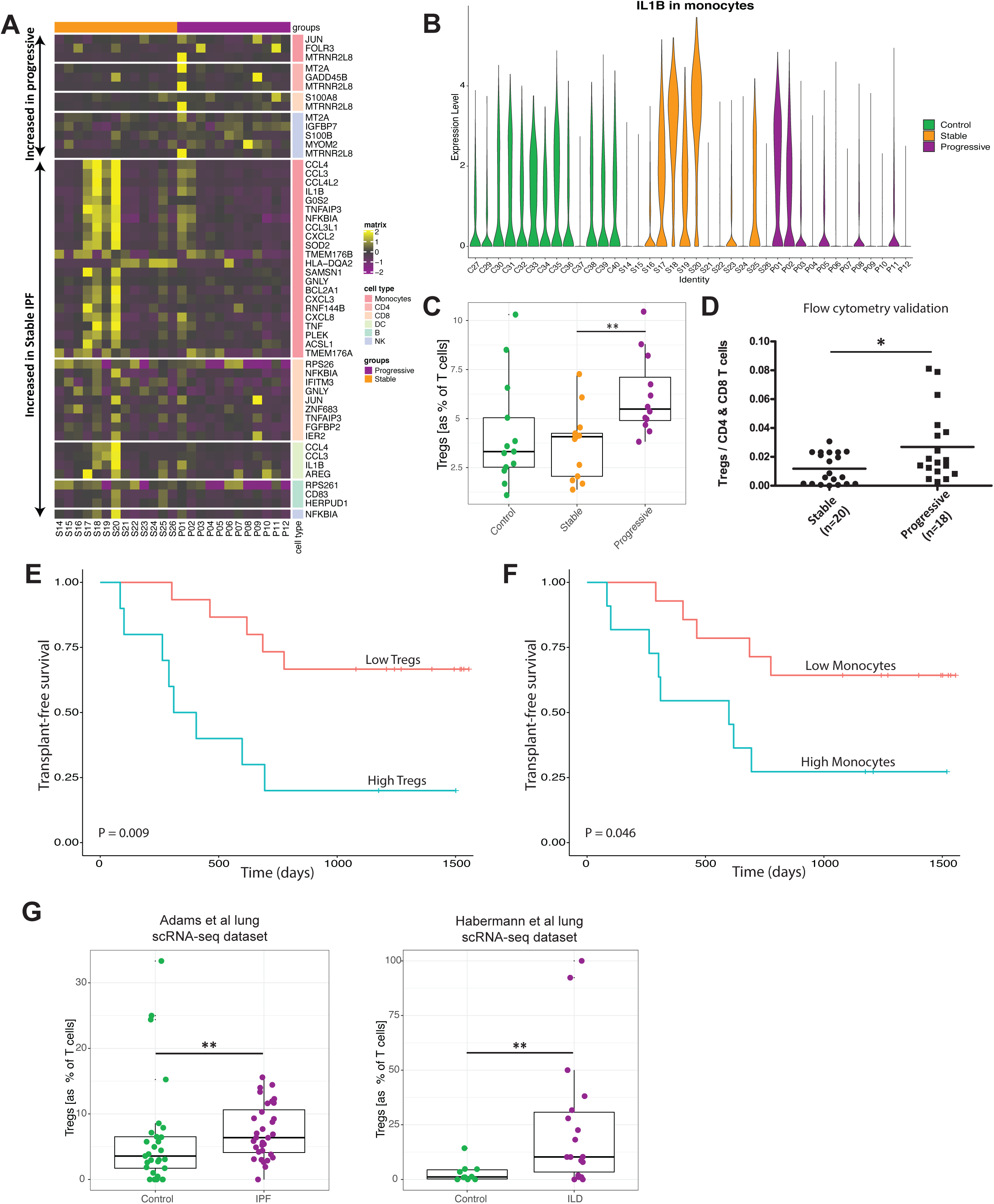
Gene expression changes in stable vs progressive IPF, and the increased Tregs in blood and lung in progressive disease. A. Heatmap showing the top differentially expressed genes (log2FC > 0.3, adjusted P values < 0.05) for each major cell type, comparing stable and progressive IPF patients. The upper part of the heatmap enumerates genes that are increased in progressive compared to stable IPF (marked “increased in progressive”), while the lower part lists genes increased in stable IPF. Each row is a gene, each column is a patient. Note the color codes for IPF severity on top and cell type on the right. B. Expression of *IL1B* in monocytes of all subjects. On average, *IL1B* expression is higher in stable compared to progressive patients. C. Box plot showing an increased level of Tregs (presented as % of all T cells) in progressive IPF subjects. Each dot represents an individual subject. D. Flow cytometry validation in an independent cohort of IPF patients, demonstrating an increase in Tregs in progressive IPF (P=0.039). E. Kaplan-Meier survival curves in IPF patients, showing a clear split of the curves based on the scRNA-seq levels of Tregs (cutoff for high Tregs [as % of T cells] >5%, P=0.009). F. Same as D but split according to the scRNA-seq levels of all monocytes (cutoff for high monocytes >39% of all PBMCs, P=0.046). G. Tregs are increased in progressive IPF/ILD in two lung scRNA-seq datasets by Adams et al (18) and Habermann et al (17), following the removal of outliers. Abbreviations: DC dendritic cells, ILD interstitial lung disease, IPF idiopathic pulmonary fibrosis, NK natural killer cells. * p<0.05, ** p<0.01.

TNFα and IL1β pathways (*TNF*, *IL1B*, *NFKBIA*, *CXCL2/3/8*, *CCL4*, *TNFAIP3*, *SOD2*) were increased in monocytes of stable IPF patients compared to progressive ones, suggesting that at least in some stable patients there is a pro-inflammatory state in monocytes (Figure 4A-B). TNFα and IL1β pathways were also increased in CD8^+^ T cells in stable IPF patients, together with higher expression of granulysin (*GNLY*) which encodes a cytolytic and proinflammatory protein present in cytotoxic granules of cytotoxic T lymphocytes and NK cells (28). Several MHC class II molecules were increased in NK cells in stable patients (Table E3), indicating a higher level of NK activation (29) in those patients. Concordantly, the composite score for all MHC class II molecules (HLA2 score) was higher in NK cells from stable IPF patients, especially in NK CD56^bright^ subpopulation (Figure E6).

Together, these gene expression changes in the peripheral blood immune system indicate a more activated pro-inflammatory state in stable IPF patients compared to those with progressive disease.

### Regulatory T cells are increased in progressive disease, in both blood and lung

Tregs were increased in the peripheral blood of progressive vs stable IPF patients, both as percentage of all PBMCs (1.8% vs 1.1%, P=0.007; Figure 2B) and as % of all T cells (6.2% vs 3.7%, P=0.003; Figure 4C). This increase is even more striking considering that the total lymphocyte count decreases in progressive disease (Figure 2A). We validated this finding in an independent cohort of IPF patients, using an existing flow cytometry dataset (Tregs as percentage of CD4+ and CD8+ T cells: 2.7% vs 1.2%, P=0.039; Figure 4D). Furthermore, Tregs levels may have a prognostic significance: survival curves clearly diverged with significantly higher mortality in patients with higher Treg counts, despite our relatively low sample size (P=0.009; Figure 4E). This resembled the survival curves for monocyte levels, which are known to be prognostic (8) (P=0.046; Figure 4F). Following this observation, we wished to explore whether Tregs are increased in the lungs as well. We re-analyzed the lung scRNA-seq datasets of Adams et al (18) and Habermann et al (17) and found increased Tregs in progressive fibrotic lungs compared to control lungs (Figure 4G). Genes belonging to Treg activation pathways (TGFβ/SMAD, mTOR and IL-2 pathways) were increased in peripheral Tregs of progressive patients, while IFNγ pathway was decreased (Figure E7), although these differences were small, and did not reach statistical significance level in our small number of Tregs (N=913 for progressive and N=604 for stable). Taken together, these findings suggest that in progressive IPF patients, activated Tregs may be recruited from the blood into the diseased lungs, and are of prognostic relevance.

### Combined evidence from lung and blood associates specific cytokines with recruitment of monocytes and Tregs in IPF

Next, we aimed to uncover specific cytokines and chemokines that may play a role in monocyte or Treg accumulation in IPF. Plasma samples were obtained from the same blood draw as the PBMC samples. For monocytes we measured plasma levels of the colony-stimulating factors (M-CSF, G-CSF, GM-CSF) and monocyte chemoattractant proteins (MCPs: CCL2, CCL7, CCL8, CCL13), which are ligands of CCR2, a major chemoattractant receptor on monocytes (30) (Figure 5A). For Tregs, based on our scRNA-seq data showing high specificity of *CCR4* and *CCR8* receptors to Tregs (Figure 5A), we measured plasma levels of their ligands (CCL17 / CCL22 and CCL1 / CCL18, respectively).

**Figure 5:**
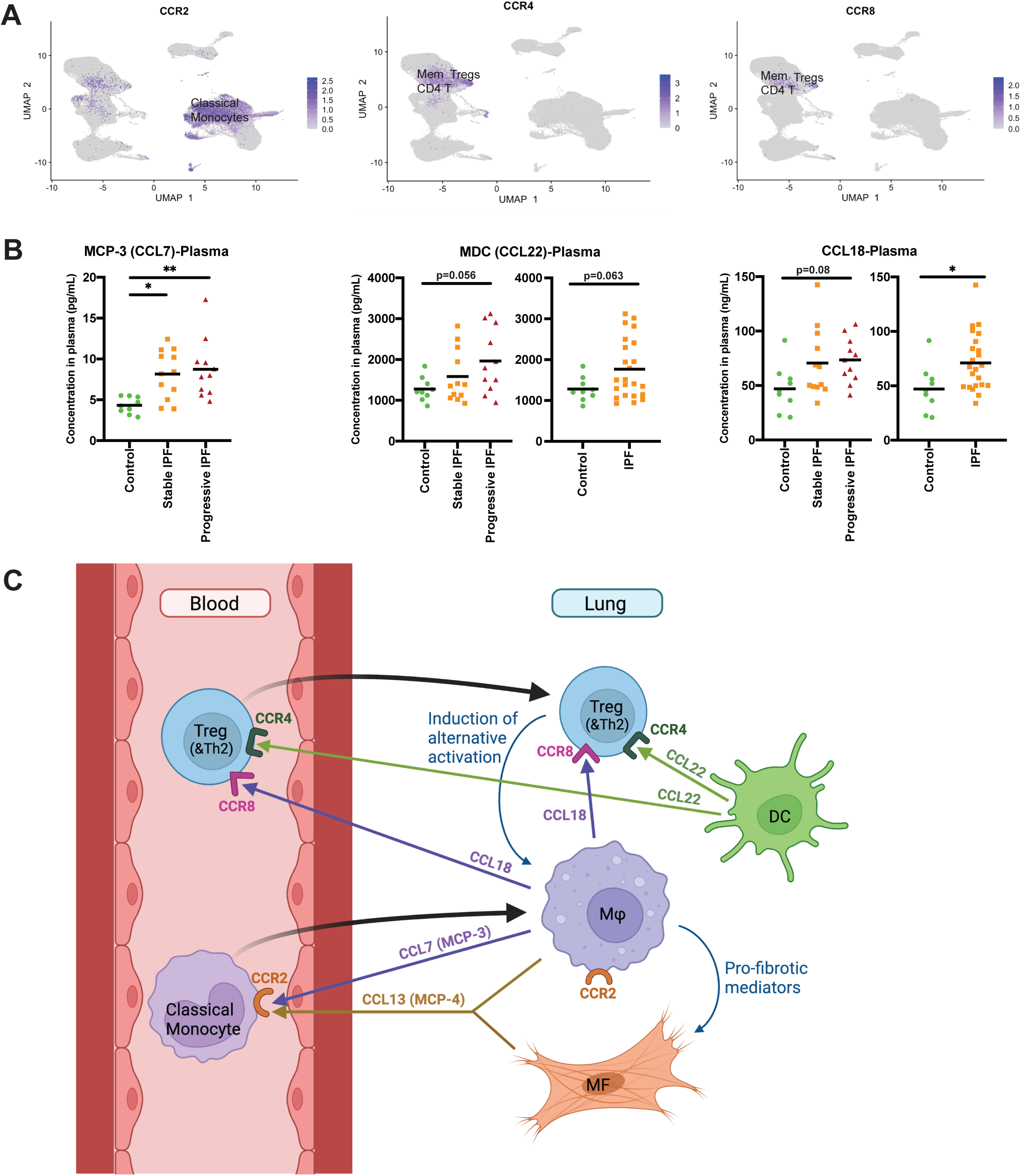
Potential chemoattractants of monocytes and Tregs in IPF and a proposed lung-blood recruitment model. A. Expression of the monocyte chemokine receptor *CCR2*, and of the Treg chemokine receptors *CCR4* & *CCR8* in IPF PBMCs. Note that *CCR4* & *CCR8* are also expressed in memory CD4 T cells, especially Th2 cells (see Supplementary Figure E8), and that non-classical monocytes lack the expression of CCR2. B. Chemokine ligands (CCL7, CCL22, CCL18) of the above respective chemokine receptors (CCR2, CCR4, CCR8) that were increased in plasma of IPF patients (N=12 stable, and 11 progressive) compared to controls (N=9). Black bars indicate the mean values. These chemokines are potential candidates that may be involved in chemoattraction of classical monocytes and Tregs from the blood into the IPF lung. Supplementary Figure E9 contains levels of other cytokines and chemokines measured in plasma and lung tissue homogenates. C. Proposed integrative lung-blood recruitment model in IPF. The left part of the figure depicts Tregs and classical monocytes in the peripheral blood, while the right part is dedicated to lung cells. Lung macrophages (Mϕ) and myofibroblasts (MF) secrete CCL7 (and potentially CCL13) into the blood, which stimulate CCR2-mediated recruitment of classical monocytes. These monocytes migrate to the IPF lung (black arrow) and are thought to be precursors of lung macrophages. Lung macrophages and dendritic cells (DC) secrete CCL18 and CCL22, respectively. These chemokines may drive CCR8 and CCR4-mediated recruitment of Tregs and Th2 cells, which may migrate to the fibrotic niche and induce a pro-fibrotic secretory profile in macrophages. Figure 5C was created with BioRender.com. * p<0.05, ** p<0.01.

Figure 5B highlights the three main chemokines that were found to be increased in IPF plasma. Among the four MCPs, CCL7 (MCP-3) showed a significant increase in IPF plasma compared to control levels, both in stable (P<0.05) and progressive patients (P<0.01). While CCL13 (MCP-4) levels did not change in IPF plasma, they were significantly increased in IPF lung tissue homogenates (P<0.001; Figure E9). As for CCR4 ligands, plasma CCL22 levels showed a trend towards an increase, both in IPF vs controls (P=0.063) and specifically in progressive IPF vs controls (P=0.056), while CCL17 did not significantly change. CCL22 levels were significantly increased in IPF lung tissue homogenates compared to controls (P<0.001; Figure E9). CCR8 ligands include CCL1 which was not elevated in IPF plasma, and CCL18, a well-known blood biomarker that predicts outcome in IPF (31), which we confirmed was elevated in IPF compared to controls (P<0.05). M-CSF, G-CSF and GM-CSF plasma levels were not changed between the three patient subgroups as shown in Figure E9, together with the results for the other cytokines tested in plasma and lung tissue homogenates.

### Lung-blood recruitment model in IPF

In the previous sections we demonstrated that monocytes and Tregs are increased in progressive IPF, and that specific chemokines may play a role in their recruitment to the lung. Next, we used the lung scRNA-seq datasets of Adams et al (18) and Habermann et al (17) to determine the cellular origin of the above chemokines in the IPF lung (Figure E10). *CCL22* is mainly expressed by dendritic cells in the lung. Macrophages have the highest level of *CCL18* expression. *CCL7* is mainly expressed by macrophages, while *CCL13* by macrophages and myofibroblasts. Based on these findings, we propose a lung-blood recruitment model in IPF (Figure 5C), that will be elaborated in the discussion section below.

### Monocyte and lymphocyte shifts drive the outcome-predictive 52-gene signature

The 52-gene signature is a validated “bulk” RNA sequencing biomarker predictive of outcome in IPF patients (6, 7), comprised of 7 genes that increase and 45 genes that decrease in association with shorter transplant-free survival (TFS). Looking into the cellular origin of the 52-gene signature, 50 of the 52 genes were detected in our dataset (7/7 and 43/45 genes, respectively). The 7 increased genes that correlate with shorter TFS were mainly of monocyte origin, while the 43 decreased genes originated mainly from T, B and NK cells (Figure 6), suggesting that the 52-gene signature is driven, at least in part, by the increase in monocytes and decrease in lymphocytes observed in progressive vs stable IPF.

**Figure 6:**
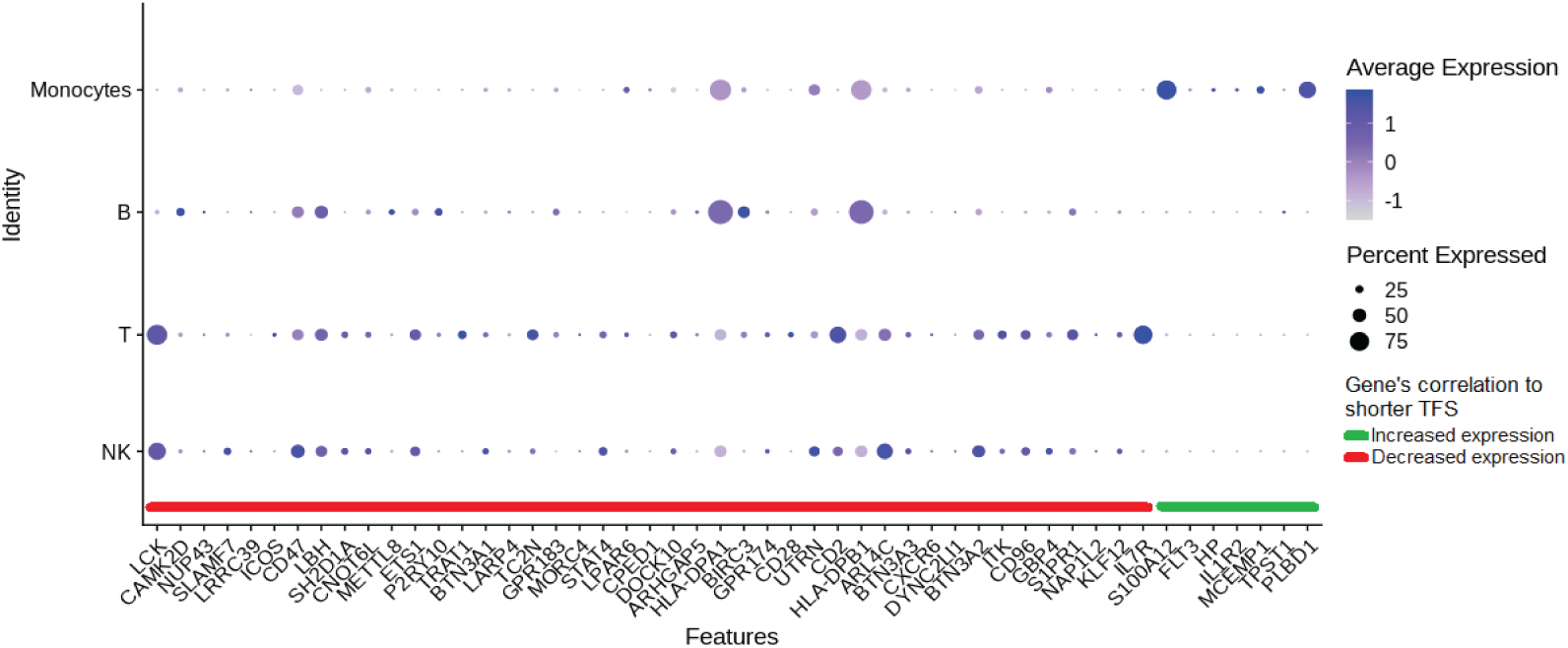
Cellular origin of the outcome-predictive 52-gene signature. Dot plot showing the cellular origin of the 52-gene signature in our scRNA-seq cohort. Each row represents a binned cell type, while each column represents a gene. Dot size indicates the percentage of cells expressing a gene, while its color intensity represents the average expression. Fifty of the 52 genes were detected in our dataset. The 7 increased genes that correlate with shorter TFS (green) were mainly of monocyte origin, while the 43 decreased genes (red) originated mainly from T, B and NK cells. Abbreviations: NK natural killer, TFS transplant-free survival.

## Discussion

In this study, we provide a comprehensive atlas of the peripheral immune system in IPF according to disease progression. We demonstrate that classical and intermediate monocytes, but not non-classical ones, are increased in progressive IPF, are associated with increased mortality, and differ in their inflammatory properties between stable and progressive disease. While many lymphocyte populations are decreased in progressive disease including naïve and effector T cells, Tregs are increased in the peripheral blood in progressive IPF and are associated with decreased survival. Increased Tregs are also found in two cohorts of lung scRNA-seq. Using our PBMC scRNA-seq dataset, we identify *CCR4* and *CCR8* as chemokine receptors potentially specific to Tregs and measure their ligands (CCL22 and CCL18, respectively) in plasma and lung tissue homogenates. CCL22, a chemokine secreted by lung DCs, is increased both in IPF plasma and IPF lung tissue homogenates, while CCL18 is only increased in IPF plasma. Additionally, CCL7, a ligand of the monocyte chemokine receptor CCR2, is increased in the plasma of IPF patients. Based on our findings, we propose a lung-blood recruitment model for monocytes and Tregs in IPF.

Our combined blood and lung scRNA-seq data, validated by flow cytometry, suggest that Tregs are increased in progressive IPF and are associated with increased mortality. The role of Tregs in IPF has previously been studied, yielding conflicting results (2, 32). An initial report that demonstrated reduced quantities and function of Tregs in peripheral blood of patients with IPF compared to healthy controls (33), has been followed by several studies showing the opposite (21, 34). Our results may resolve this disagreement; we observed a decrease in Tregs in stable disease compared to controls, but an increase in Tregs in progressive vs stable IPF, which associates with worst outcomes. Thus, it is possible that previous work differed in the admixture of progressive and stable IPF and highlights the need to study these groups independently.

To better understand what drives Tregs to the lung we explored receptors and ligands potentially involved in Treg migration in IPF. Using our scRNAseq data we found that *CCR4* and *CCR8* are specific chemokine receptors expressed mainly on Tregs, and may therefore explain selective Treg migration while most other lymphocyte subpopulations are attenuated. Previous reports have shown that Tregs express more *CCR4* than other T cells (35) and that anti-CCR4 monoclonal antibodies selectively deplete Tregs from human tissues (36). We demonstrate that CCL22, a CCR4 ligand, is increased both in IPF plasma and IPF lung tissue homogenates, and thus is a likely candidate to be involved in Treg chemoattraction in IPF. This conclusion is further strengthened by previous studies linking CCL22 to Treg trafficking. Mailloux et al reported that CCL22 secreted from lung tissue selectively attracted Tregs (35), and Wei et al found that CCL22 mediated Treg cell trafficking into human ovarian cancer via CCR4 (37). CCL22 was also found to be increased in BAL fluid and its levels inversely correlated with DLco/VA (38). Although CCL17, the other CCR4 ligand, was not increased in our plasma samples or lung tissue homogenates, we cannot exclude its involvement in IPF. Like CCL22, CCL17 was found to be increased in BAL fluid (38), and more recently a large-scale plasma proteomic profiling study identified CCL17 as one of the proteins with the greatest difference in IPF versus controls (39). Similar to CCR4, CCR8 receptor may also be involved in Treg migration, and was previously shown to be expressed mainly on Tregs and Th2 cells (40). CCL18, a well-recognized prognostic blood biomarker in IPF (31), is a ligand for CCR8 receptor (41), and was previously demonstrated to recruit human Tregs (42). CCL18 is secreted by lung macrophages and was found to be elevated in our IPF vs control plasma.

Blood monocytes are thought to be the precursors of lung macrophages, which were shown to play an important pro-fibrotic role in the fibrotic niche (2, 43). Our scRNA-seq data shows that increased blood monocytes are associated with decreased survival (8, 9), and that this increase drives, at least in part, the prognostic 52-gene signature. CCR2 is a major chemoattractant receptor on monocytes (30) and CCR2 knockout mice were protected from developing pulmonary fibrosis (44). We demonstrate that the levels of classical and intermediate monocytes increase in IPF blood, especially in progressive disease, and that the CCR2 ligands CCL7 (and possibly CCL13) are increased in IPF plasma (or lung tissue homogenates, respectively) and thus may play a role in monocyte recruitment into the IPF lung. Interestingly, non-classical monocytes are the only monocyte subpopulation that did not increase in IPF, potentially related to their lack of expression of CCR2 (Figure 5A). CCL7, also known as MCP-3, was previously shown to be increased in surgical lung biopsies from patients with usual interstitial pneumonia (UIP) compared to other idiopathic interstitial pneumonias (IIP) and patients without IIP (45). CCL7 and CCL2 are considered the CCR2 agonists that are most critical for monocyte mobilization (30). Since a phase II clinical trial of anti-CCL2 monoclonal antibodies in IPF was negative (46), we believe that the role of CCL7 in IPF merits further investigation. Evidence regarding the role of CCL13 in IPF is scarce.

Based on our blood and lung gene expression and protein findings, we propose a lung-blood recruitment model in IPF (Figure 5C). Dendritic cells and macrophages recruit Tregs (and possibly Th2 cells) into the fibrotic IPF lung by secreting CCL22 and CCL18, which serve as ligands for their chemokine receptors CCR4 and CCR8. Tregs were previously shown to induce an alternative activation phenotype in macrophages (47), and Th2 cells are part of the Th2 response, both thought to play a pro-fibrotic role in IPF (2). Macrophages and myofibroblasts secrete the monocyte chemoattractants CCL7 and CCL13, thus recruiting classical monocytes to serve as precursors to the pro-fibrotic IPF macrophages. We emphasize that this model needs to be further validated in future studies.

Our study has several limitations. Although it is a relatively large scRNA-seq study with 38 samples, the number of samples in each of the three subgroups (stable IPF, progressive IPF and control subjects) is relatively small. Nevertheless, numerous statistically significant findings were detected. In addition, we had to focus on a limited number of chemokines and cytokines to be measured in the plasma, and therefore cannot exclude a role for other secreted agents in the accumulation of immune cells in IPF. Lastly our lung-blood model requires validation in future studies.

In summary, we provide a first single-cell atlas of the peripheral immune system in stable and progressive IPF. The results of our peripheral blood scRNA-seq data, integrated with our previous IPF lung scRNA-seq and protein findings, reveal an outcome-predictive increase in classical monocytes and Tregs in IPF, suggesting the presence of an IPF lung-blood immune recruitment axis involving CCL7 (for classical monocytes) and CCL18/CCL22 (for Tregs). Further study of this novel lung-blood immune recruitment model may yield insight into IPF and related diseases, as well as potential novel immune interventions in this devastating disease.

## Data Availability

Results can be further explored through our user-friendly data-mining website (http://ildimmunecellatlas.com), and raw sequencing data will be available in GEO upon peer-reviewed publication.

## Acknowledgment

We are indebted to all patients and control subjects who participated in this study. The sequencing was conducted by Mei Zhong and her team at Yale Stem Cell Center Genomics Core facility. We wish to thank Taylor Adams and John McDonough for their helpful comments and advice, Xue Yan Peng for her help in processing the samples for CyTOF, and Julia Winkler for biobanking the PBMC samples.

## List of Supplementary Information

### Supplementary Figures

E1 Results of the unsupervised clustering and cell type annotation markers

E2 Automated annotation of cell types with SingleR package

E3 CyTOF validation of cell counts

E4 COL1A1+CD45+ cells

E5 Platelet-monocyte complexes

E6 Pathway analysis of DEGs in stable vs progressive PBMC subpopulations

E7 Pathways altered in Tregs of progressive vs stable IPF

E8 Distribution of *CCR4* and *CCR8* receptors on PBMCs

E9 Plasma and tissue homogenate levels of cytokines and chemokines

E10 Cellular origin of selected cytokines in the IPF lung, based on scRNA-seq datasets

### Supplementary tables

E1 Randomization of samples into five batches

E2 Differentially expressed genes in IPF patients vs controls

E3 Differentially expressed genes in stable vs progressive IPF

E4 List of antibodies used in CyTOF

