## Supplementary Figure E1 for "Single-cell profiling reveals immune aberrations in progressive idiopathic pulmonary fibrosis"

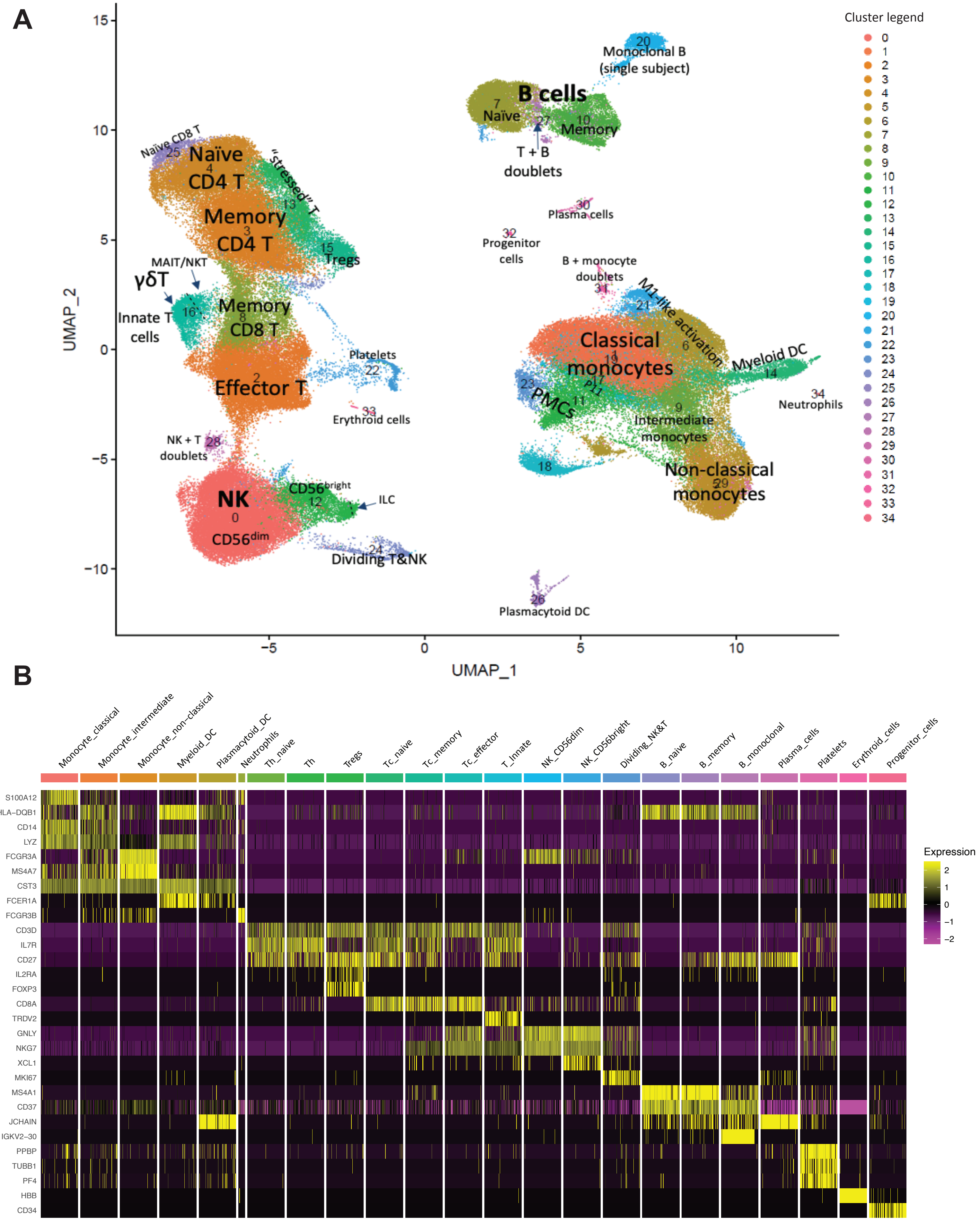


**Figure E1: Results of the unsupervised clustering and cell type annotation markers.** A. Preliminary UMAP of 153,162 cells, showing 35 clusters with detailed cell-type annotation. Five doublet clusters (#19, 27, 28, 29, 31) were removed, and the final Seurat object for analysis consisted of 149,564 cells across 30 clusters aggregated into 23 cell types (as shown in Figure 1B). B. Heatmap showing the expression of specific cell type annotation markers used for manual annotation, across all cell types.
