## Supplementary Figure E2 for "Single-cell profiling reveals immune aberrations in progressive idiopathic pulmonary fibrosis"

A

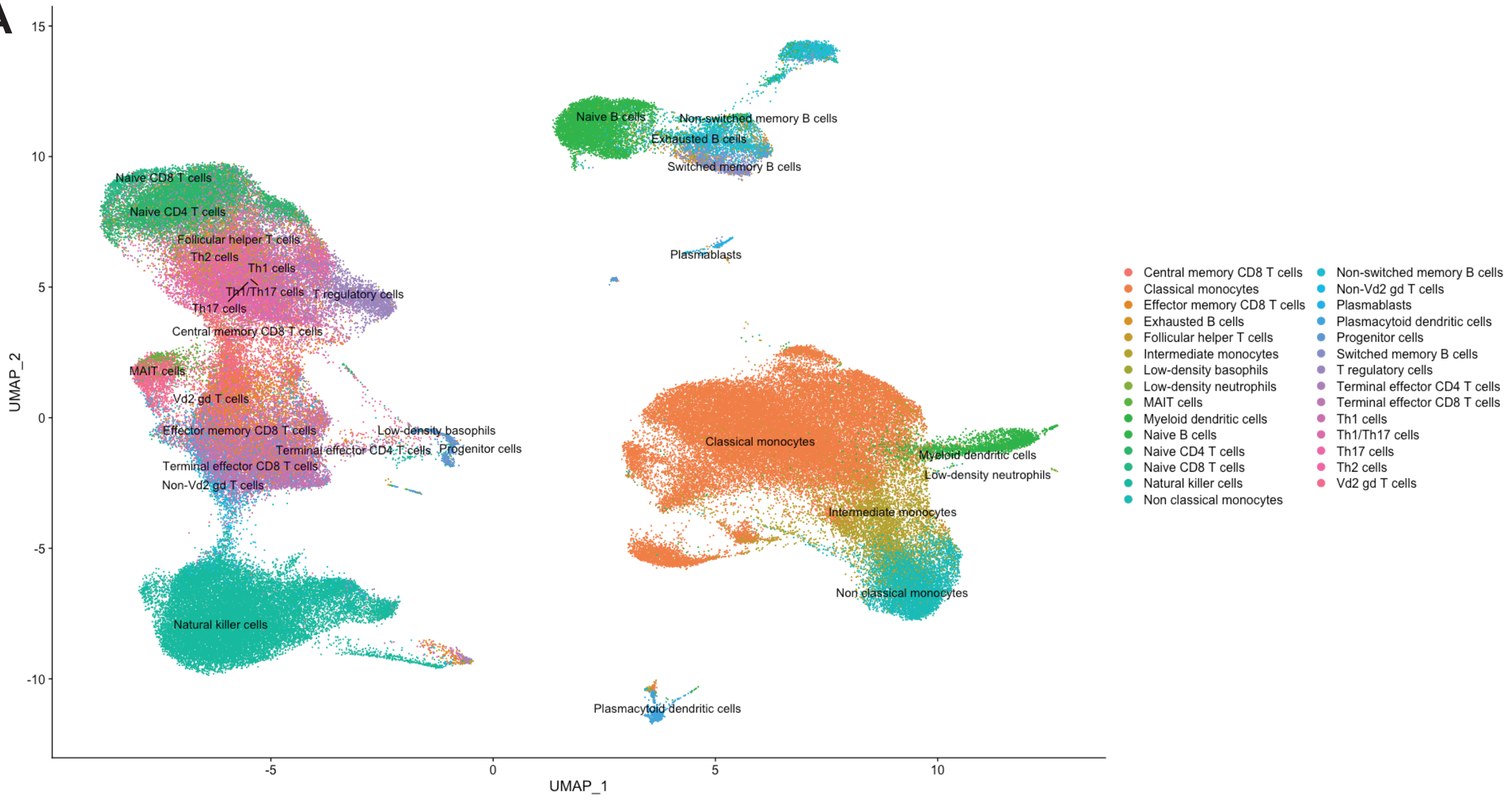


B

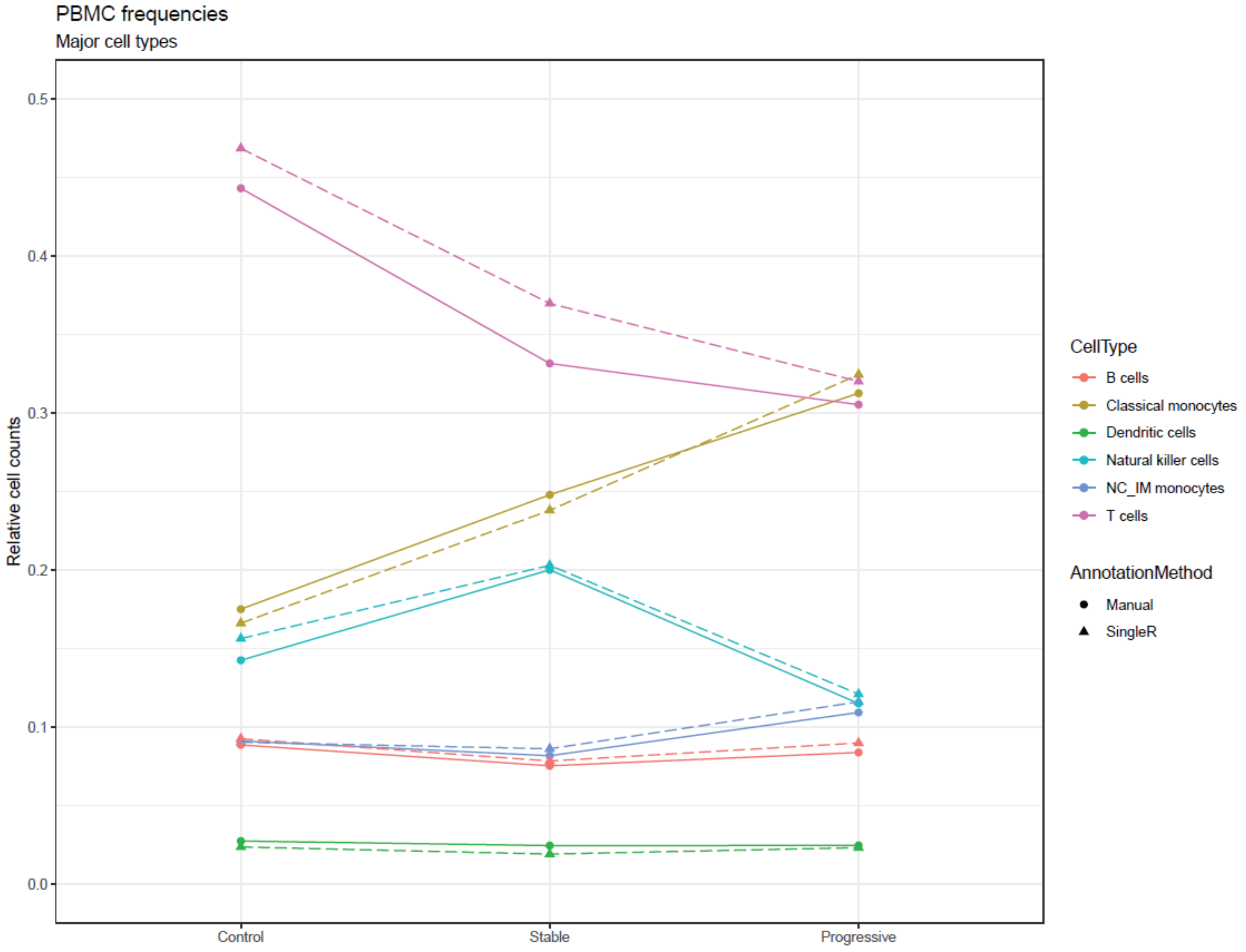


**Figure E2: Automated annotation of cell types with SingleR package.** A. UMAP showing automated annotation of cells into 29 cell types, using SingleR package. Please refer to the UMAP with manual annotation in Figure 1B. B. Comparison of manual (circles) and automated (triangles) annotation results for several major cell types, showing similar cell frequencies across all three patient subgroups. Lines are given for convenience only, in order to highlight this similarity.
