## Supplementary Figure E3 for "Single-cell profiling reveals immune aberrations in progressive idiopathic pulmonary fibrosis"

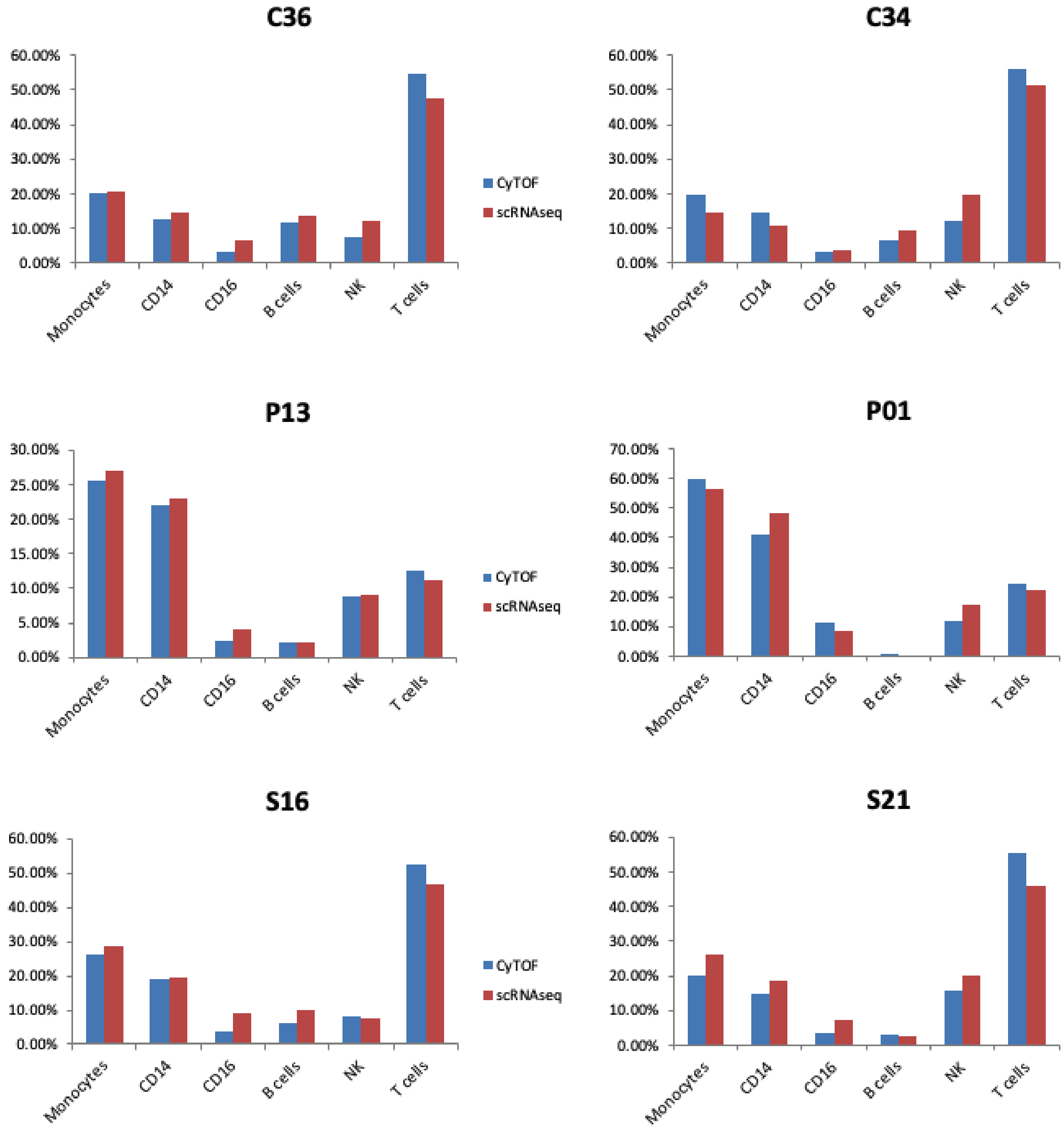


**Figure E3: CyTOF validation of cell counts.** Relative cell counts of main cell types (as % of all PBMCs) comparing scRNA-seq based counts to CyTOF-based counts. Each of these samples was split into 2 and ran in parallel using both methods. Overall, there was excellent agreement between both methods (mean correlation coefficient is 0.99, range 0.97-1.00). The total monocytes column is shown on the left, and includes CD14+/CD16- and CD16+ monocytes which are also shown in two separate columns. CD16+ population in scRNA-seq includes both intermediate and non-classical monocytes clusters. P13 sample was excluded from the full scRNA-seq analysis due to low sample quality, but was still used here for cell count validation. See Supplementary Methods section for more details on our CyTOF methodology.
