## Supplementary Figure E4 for "Single-cell profiling reveals immune aberrations in progressive idiopathic pulmonary fibrosis"

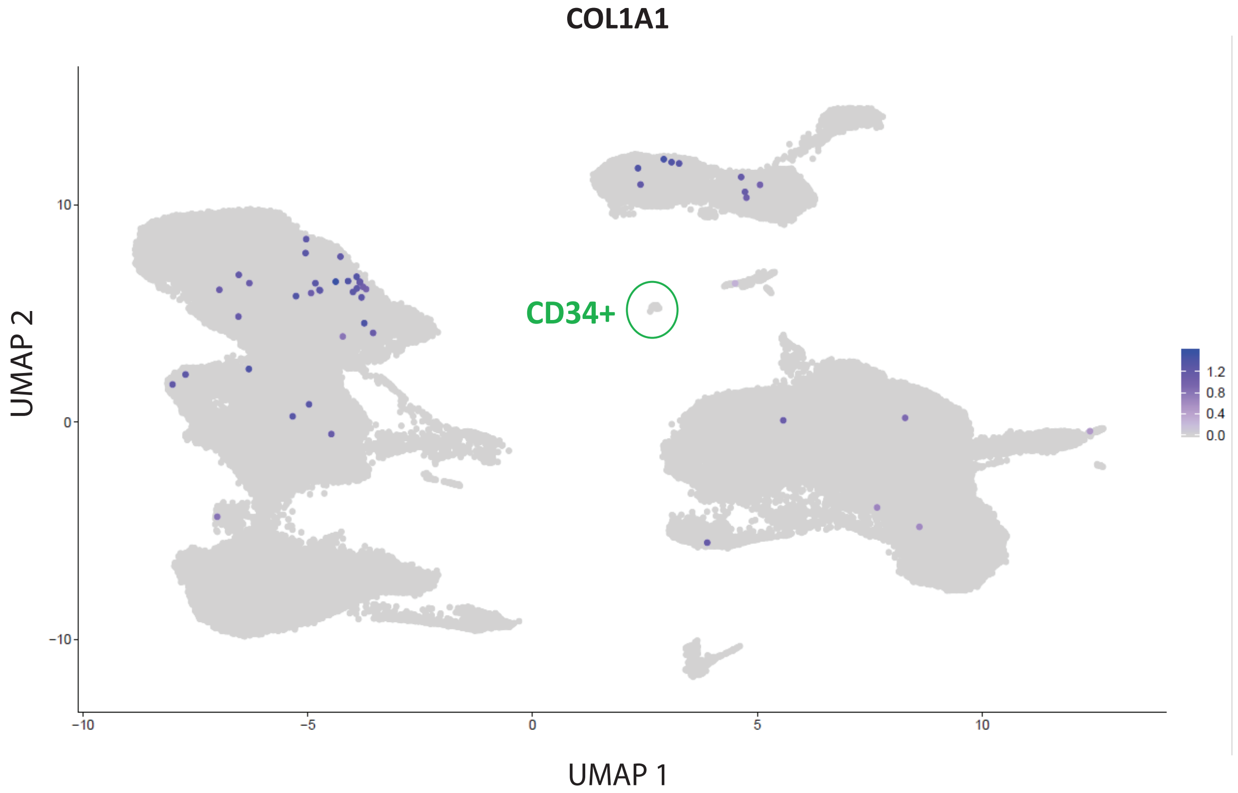


**Figure E4: COL1A1+CD45+ cells.** UMAP highlighting all cells that express *COL1A1* (and co-express *CD45)*, which are fibrocyte markers. Those cells were rarely found in our dataset (0.03% of all cells), were scattered, and did not form their own cluster. None of them were found among the CD34+ progenitor cells cluster (in green circle).
