## Supplementary Figure E5 for "Single-cell profiling reveals immune aberrations in progressive idiopathic pulmonary fibrosis"

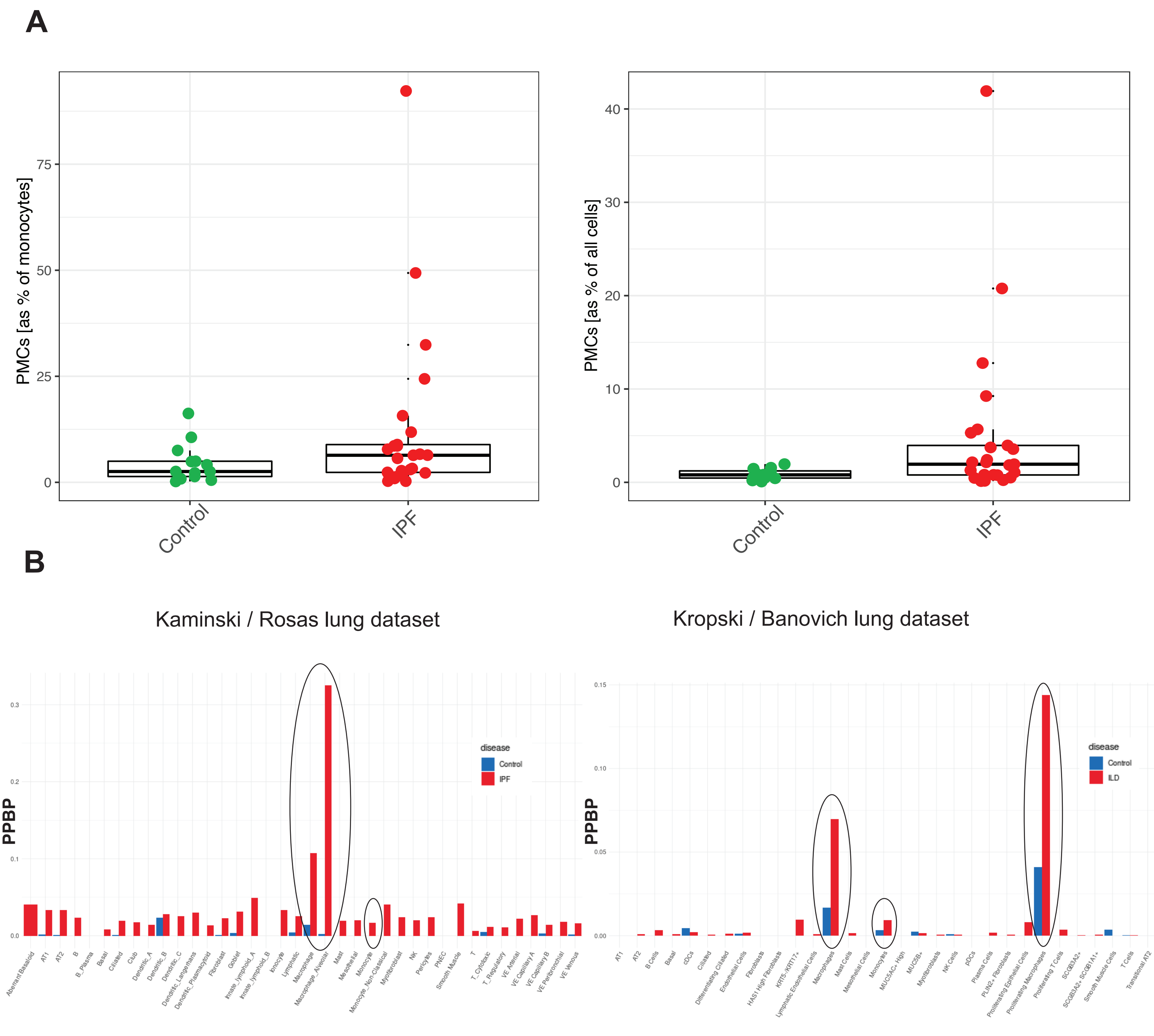


**Figure E5: Platelet-monocyte complexes.** A. Boxplots comparing platelet-monocyte complexes (PMCs) in IPF patients vs control subjects. Two clusters (#11, 23) of platelet-monocyte complexes (PMCs) were identified in our data. The number of cells in these PMC clusters (as percentage of all cells) was higher in IPF patients compared to control subjects (p=0.03). B. Levels of *PPBP* were found to be increased in pulmonary fibrosis vs control monocytes and macrophages in two datasets of lung scRNA-seq.
