## Supplementary Figure E6 for "Single-cell profiling reveals immune aberrations in progressive idiopathic pulmonary fibrosis"

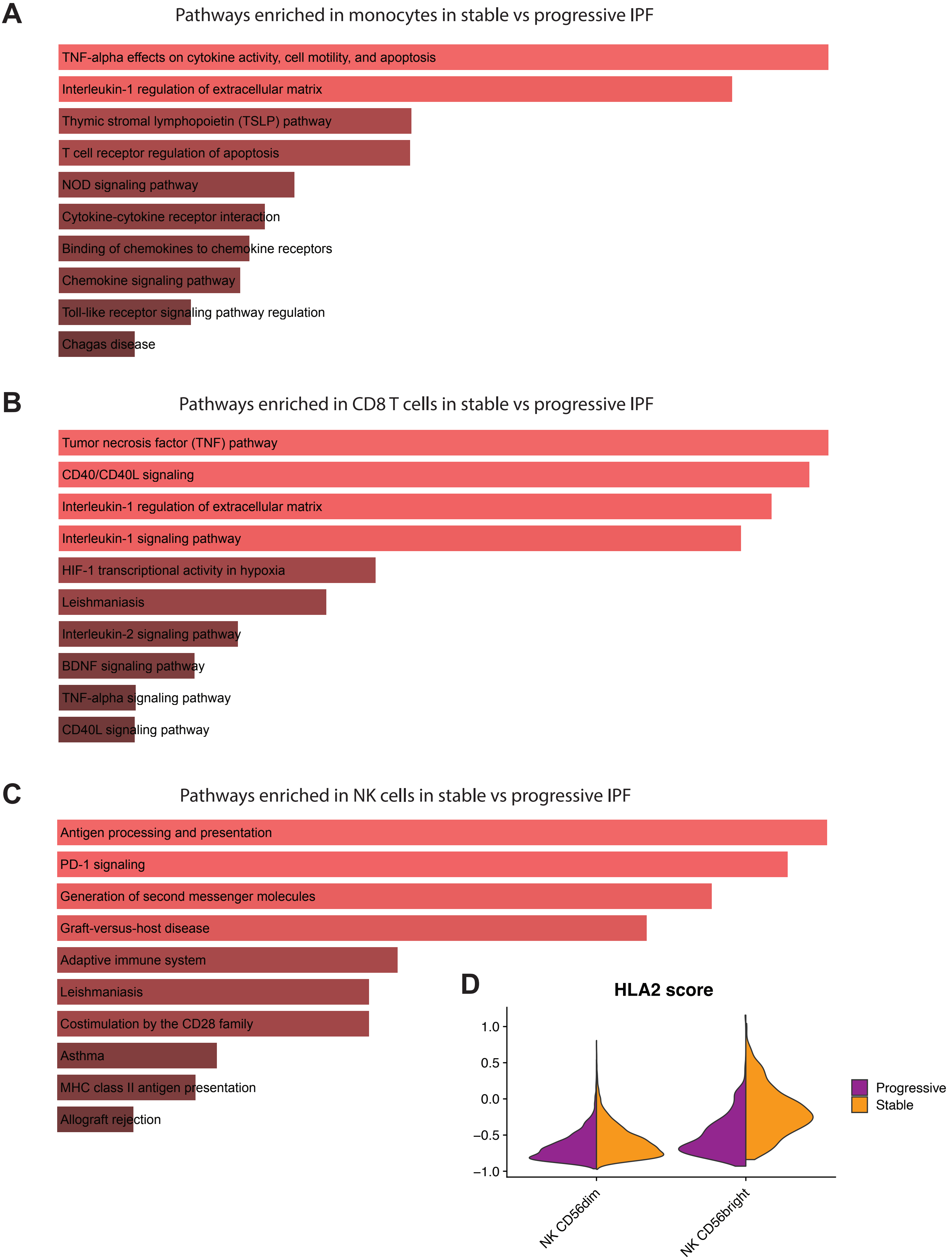


**Figure E6: Pathway analysis of DEGs in stable vs progressive PBMC subpopulations.** A-C. Pathway enrichment analysis of differentially expressed genes (DEGs) in stable vs progressive IPF, conducted for monocytes, CD8+ T cells, and NK cells, respectively. These analyses were conducted using the Enrichr online platform (available at https://maayanlab.cloud/Enrichr/). D. Violin plot showing higher expression of HLA class II molecules in NK cells of stable IPF patients, especially the NK CD56bright subpopulation. This is concordant with the enriched pathway of antigen processing and presentation shown in figure C.
