## Supplementary Figure E7 for "Single-cell profiling reveals immune aberrations in progressive idiopathic pulmonary fibrosis"

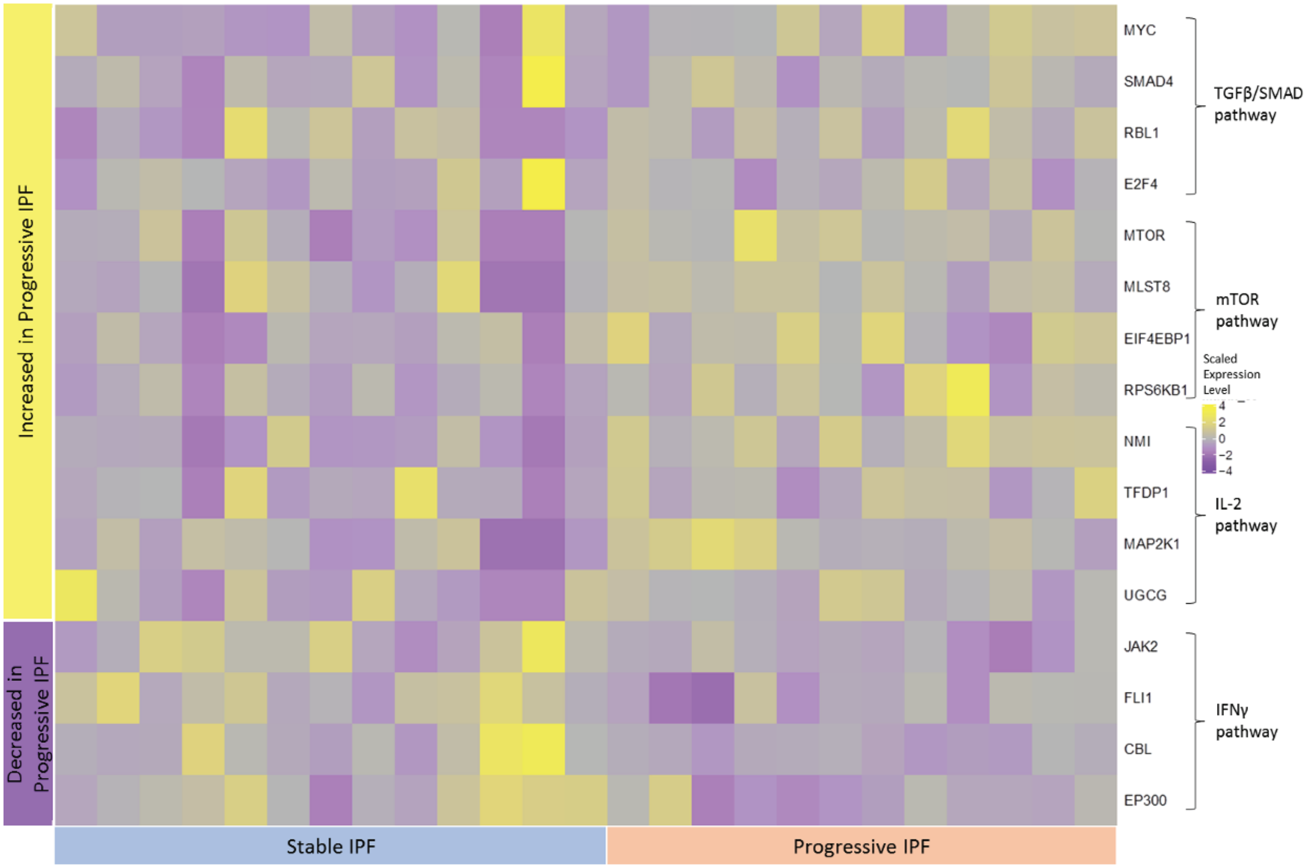


**Figure E7: Pathways altered in Tregs of progressive vs stable IPF.** Heatmap showing selected genes/pathways in Tregs. Each row represents a gene and each column a patient. Genes belonging to Treg activation pathways (TGFβ/SMAD, mTOR and IL-2 pathways) were increased in progressive patients, while those of IFNγ pathway were decreased. Of note, these differences were small, and we did not reach statistical significance level in our small cohort of Tregs (N=913 for progressive and N=604 for stable).
