## Supplementary Figure E8 for "Single-cell profiling reveals immune aberrations in progressive idiopathic pulmonary fibrosis"

### A *CCR4* receptors on PBMCs

| | | | | | |
|---|---|---|---|---|---|
| Central memory CD8 T cells | 93 | Classical monocytes | 30 | Effector memory CD8 T cells | 75 |
| Exhausted B cells | 0 | Follicular helper T cells | 151 | Intermediate monocytes | 13 |
| Low-density basophils | 1 | Low-density neutrophils | 0 | MAIT cells | 1 |
| Myeloid dendritic cells | 4 | Naive B cells | 0 | Naive CD4 T cells | 7 |
| Naive CD8 T cells | 1 | Natural killer cells | 6 | Non classical monocytes | 3 |
| Non-switched memory B cells | 2 | Non-Vd2 gd T cells | 2 | Plasmablasts | 0 |
| Plasmacytoid dendritic cells | 0 | Progenitor cells | 41 | Switched memory B cells | 6 |
| T regulatory cells | 591 | Terminal effector CD4 T cells | 3 | Terminal effector CD8 T cells | 2 |
| Th1 cells | 199 | Th1/Th17 cells | 57 | Th17 cells | 209 |
| Th2 cells | 582 | Vd2 gd T cells | 31 | | |

### B *CCR8* receptors on PBMCs

| | | | | | |
|---|---|---|---|---|---|
| Central memory CD8 T cells | 28 | Classical monocytes | 2 | Effector memory CD8 T cells | 7 |
| Exhausted B cells | 0 | Follicular helper T cells | 1 | Intermediate monocytes | 3 |
| Low-density basophils | 0 | Low-density neutrophils | 0 | MAIT cells | 0 |
| Myeloid dendritic cells | 1 | Naive B cells | 0 | Naive CD4 T cells | 0 |
| Naive CD8 T cells | 0 | Natural killer cells | 4 | Non classical monocytes | 1 |
| Non-switched memory B cells | 0 | Non-Vd2 gd T cells | 0 | Plasmablasts | 0 |
| Plasmacytoid dendritic cells | 0 | Progenitor cells | 0 | Switched memory B cells | 0 |
| T regulatory cells | 140 | Terminal effector CD4 T cells | 0 | Terminal effector CD8 T cells | 0 |
| Th1 cells | 34 | Th1/Th17 cells | 4 | Th17 cells | 36 |
| Th2 cells | 148 | Vd2 gd T cells | 1 | | |

**Figure E8: Distribution of *CCR4* and *CCR8* receptors on PBMCs.** A. Number of cells expressing *CCR4* for each cell type, as annotated by the SingleR package. B. Similarly, the number of cells expressing *CCR8* for each cell type. As evident, both *CCR4* and *CCR8* are mainly expressed in Tregs and Th2 cells. Please refer to Figure 5A for UMAPs of *CCR4* and *CCR8*.
