## Supplementary Figure E9 for "Single-cell profiling reveals immune aberrations in progressive idiopathic pulmonary fibrosis"

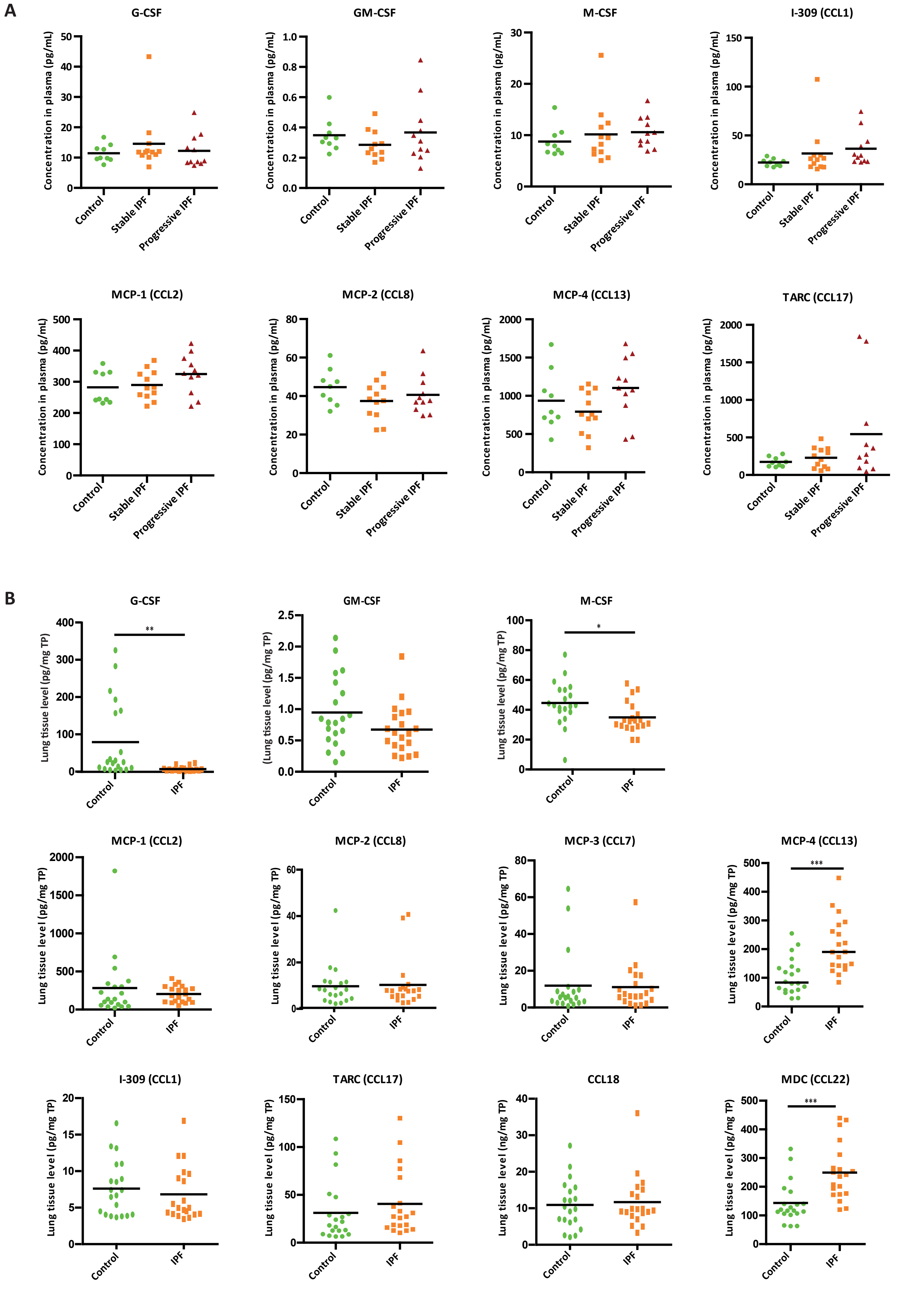


**Figure E9: Plasma and tissue homogenate levels of cytokines and chemokines.** A. Plasma levels of cytokines and chemokines, complementing those reported in Figure 5B. B. Levels of the same proteins (normalized to total protein level) in lung tissue homogenates of IPF patients (N=20) vs controls (N=20).
