## Supplementary Figure E10 for "Single-cell profiling reveals immune aberrations in progressive idiopathic pulmonary fibrosis"

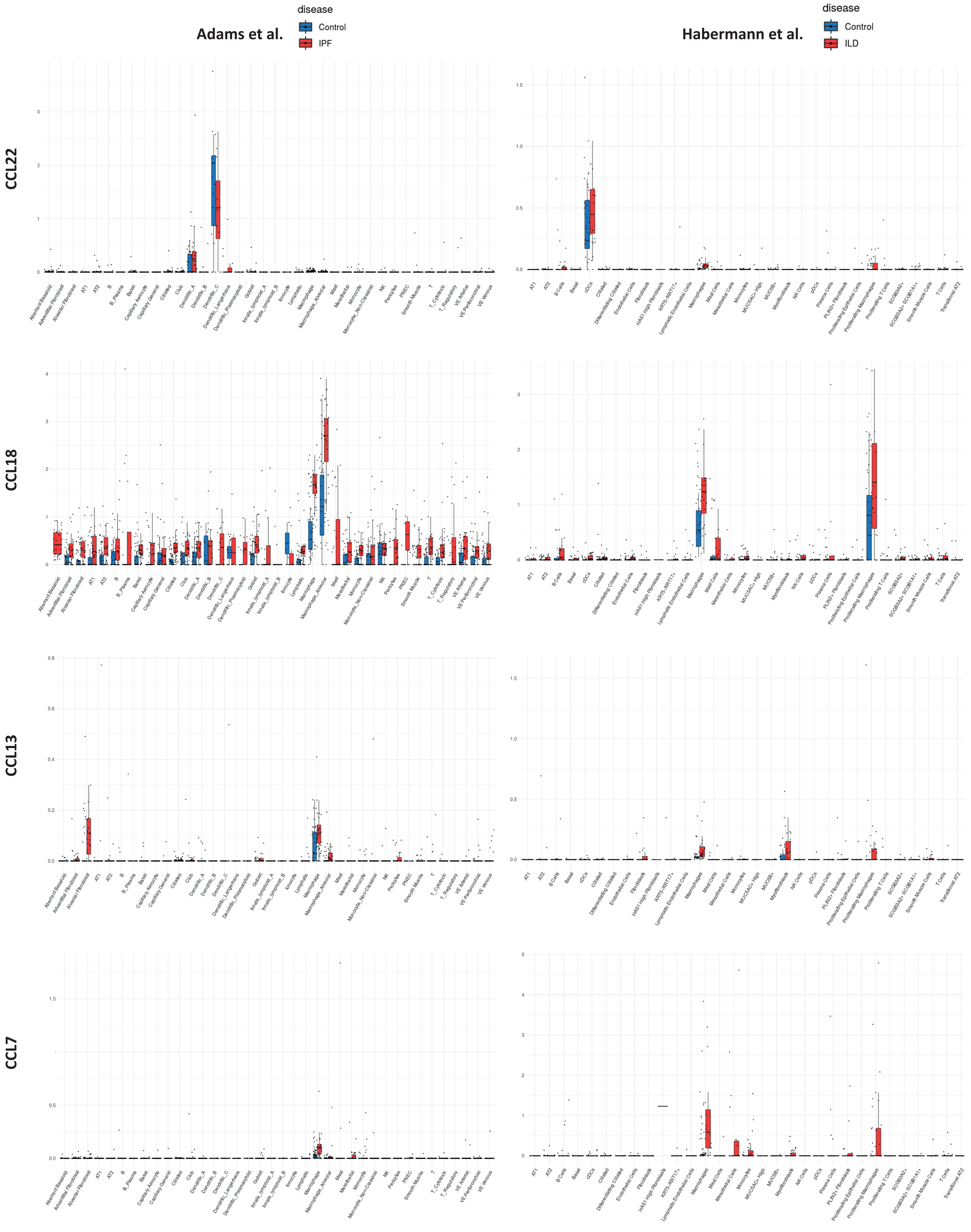


**Figure E10: Cellular origin of selected cytokines in the IPF lung, based on two scRNA-seq datasets.** Boxplots were acquired from the IPF Cell Atlas website (www.ipfcellatlas.com). Dots represent the average expression for each subject. Level of expression is shown for each cell type, with separate boxes for IPF/ILD (in red) and Control (blue). The two lung scRNA-seq datasets are Adams et al (left panels, reference 22) and Habermann et al (right panels, reference 23).
