## Supplementary Table E1 for "Single-cell profiling reveals immune aberrations in progressive idiopathic pulmonary fibrosis"

Supplementary Table E1: Randomization of samples into five batches

| Subject Code | Group assignment |
|---|---|
| **Batch 1** | |
| C36 | Control |
| C34 | Control |
| P13* | IPF- progressive |
| S21 | IPF- stable |
| C38 | Control |
| P01 | IPF- progressive |
| S23 | IPF- stable |
| S16 | IPF- stable |
| **Batch 2** | |
| S18 | IPF- stable |
| C37 | Control |
| C33 | Control |
| C29 | Control |
| P04 | IPF- progressive |
| P02 | IPF- progressive |
| S26 | IPF- stable |
| P08 | IPF- progressive |
| **Batch 3** | |
| C28* | Control |
| C32 | Control |
| P10 | IPF- progressive |
| S22 | IPF- stable |
| P03 | IPF- progressive |
| P07 | IPF- progressive |
| S17 | IPF- stable |
| S15 | IPF- stable |
| **Batch 4** | |
| P09 | IPF- progressive |
| C30 | Control |
| P12 | IPF- progressive |
| C40 | Control |
| C35 | Control |
| P06 | IPF- progressive |
| S20 | IPF- stable |
| S24 | IPF- stable |
| **Batch 5** | |
| S25 | IPF- stable |
| C31 | Control |
| S19 | IPF- stable |
| P11 | IPF- progressive |
| P05 | IPF- progressive |
| C39 | Control |
| C27 | Control |
| S14 | IPF- stable |

* Two samples (P13 and C28) were excluded following QC due to low sample quality
