## Supplementary Table E2 for "Single-cell profiling reveals immune aberrations in progressive idiopathic pulmonary fibrosis"

Supplementary Table E2: Differentially expressed genes in IPF patients vs controls

**Monocytes**

| | p_val | avg_logFC | IPF | Control | p_val_adj |
|---|---|---|---|---|---|
| ZFP36 | 0 | -0.2729314 | 0.908 | 0.966 | 0 |
| HLA-DRB1 | 0 | -0.3860753 | 0.919 | 0.964 | 0 |
| SGK1 | 0 | -0.4401736 | 0.434 | 0.611 | 0 |
| MTRNR2L12 | 9.60E-285 | -0.2936465 | 0.894 | 0.951 | 2.25E-280 |
| HLA-DRA | 1.89E-278 | -0.2978613 | 0.929 | 0.964 | 4.44E-274 |
| S100A12 | 5.19E-277 | 0.49142257 | 0.783 | 0.677 | 1.22E-272 |
| PLAUR | 2.05E-263 | -0.3851295 | 0.675 | 0.8 | 4.81E-259 |
| KLF4 | 2.35E-255 | -0.2664318 | 0.573 | 0.718 | 5.52E-251 |
| CTSD | 1.85E-216 | 0.30476776 | 0.682 | 0.563 | 4.35E-212 |
| TUBA1A | 3.92E-213 | -0.2551594 | 0.764 | 0.854 | 9.21E-209 |
| FOLR3 | 1.06E-204 | 0.51907197 | 0.29 | 0.16 | 2.48E-200 |
| S100A8 | 1.78E-201 | 0.38170155 | 0.936 | 0.892 | 4.18E-197 |
| BTG2 | 1.04E-185 | -0.2530855 | 0.602 | 0.731 | 2.45E-181 |
| NR4A1 | 6.86E-173 | -0.3168409 | 0.266 | 0.387 | 1.61E-168 |
| G0S2 | 1.41E-169 | -0.4007246 | 0.298 | 0.431 | 3.31E-165 |
| RGS2 | 2.53E-165 | -0.2778162 | 0.746 | 0.83 | 5.93E-161 |
| TMEM176B | 3.86E-164 | -0.2683048 | 0.451 | 0.591 | 9.05E-160 |
| PHACTR1 | 1.78E-157 | -0.2545917 | 0.284 | 0.407 | 4.17E-153 |
| FKBP5 | 4.57E-150 | 0.29342823 | 0.304 | 0.194 | 1.07E-145 |
| IFITM3 | 5.91E-150 | -0.3791588 | 0.775 | 0.856 | 1.39E-145 |
| PPBP | 1.35E-149 | 0.62934305 | 0.283 | 0.171 | 3.17E-145 |
| TUBA1B | 1.10E-148 | -0.2516131 | 0.697 | 0.78 | 2.58E-144 |
| HBEGF | 1.68E-144 | -0.2624931 | 0.13 | 0.222 | 3.94E-140 |
| IL1R2 | 9.73E-117 | 0.33953984 | 0.073 | 0.016 | 2.28E-112 |
| CD163 | 6.16E-110 | 0.2806015 | 0.316 | 0.222 | 1.44E-105 |
| MYL9 | 9.26E-105 | 0.31378792 | 0.125 | 0.055 | 2.17E-100 |
| CCL5 | 1.16E-94 | 0.30944047 | 0.222 | 0.137 | 2.72E-90 |
| FOSB | 2.40E-84 | -0.2634128 | 0.418 | 0.499 | 5.62E-80 |
| HLA-DQA1 | 4.10E-75 | -0.2598491 | 0.354 | 0.432 | 9.62E-71 |
| IFITM1 | 1.76E-53 | -0.2520122 | 0.26 | 0.328 | 4.12E-49 |
| FCGR3A | 1.12E-49 | -0.2729532 | 0.32 | 0.381 | 2.64E-45 |
| THBS1 | 8.34E-45 | 0.4297026 | 0.224 | 0.17 | 1.96E-40 |
| CCL4 | 2.75E-15 | 0.80928311 | 0.18 | 0.155 | 6.46E-11 |
| CCL4L2 | 7.44E-05 | 0.39937096 | 0.111 | 0.1 | 1 |
| CCL3 | 0.00323775 | 0.3330056 | 0.307 | 0.331 | 1 |
| TNFAIP3 | 0.01067987 | 0.27621252 | 0.399 | 0.418 | 1 |
| MTRNR2L8 | 0.71800613 | 1.13413685 | 0.228 | 0.244 | 1 |

**CD4+ T cells**

| | p_val | avg_logFC | IPF | Control | p_val_adj |
|---|---|---|---|---|---|
| MT-ATP8 | 0 | 0.37775155 | 0.962 | 0.93 | 0 |
| CISH | 2.51E-277 | 0.49056347 | 0.3 | 0.135 | 5.90E-273 |
| GIMAP4 | 4.31E-237 | 0.30693904 | 0.719 | 0.578 | 1.01E-232 |
| DUSP2 | 6.55E-235 | -0.5700166 | 0.211 | 0.363 | 1.54E-230 |
| PIM1 | 9.74E-201 | 0.30304001 | 0.614 | 0.459 | 2.29E-196 |
| NR4A2 | 7.28E-197 | -0.3422182 | 0.078 | 0.187 | 1.71E-192 |
| BTG2 | 2.02E-155 | -0.316607 | 0.38 | 0.508 | 4.73E-151 |
| DNAJB1 | 3.20E-148 | -0.2665365 | 0.583 | 0.678 | 7.50E-144 |
| ZFP36 | 3.01E-143 | -0.5433171 | 0.562 | 0.63 | 7.05E-139 |
| TNFAIP3 | 1.10E-136 | -0.299743 | 0.368 | 0.486 | 2.58E-132 |
| MTRNR2L12 | 1.74E-125 | -0.2581162 | 0.898 | 0.938 | 4.07E-121 |
| CXCR4 | 4.81E-106 | -0.2870877 | 0.534 | 0.615 | 1.13E-101 |
| JUNB | 4.43E-84 | -0.4636851 | 0.931 | 0.875 | 1.04E-79 |
| FOS | 3.40E-75 | -0.3981754 | 0.167 | 0.241 | 7.97E-71 |
| HIST1H1E | 7.30E-55 | -0.2507916 | 0.23 | 0.299 | 1.71E-50 |
| MTRNR2L8 | 0.00123963 | 0.82238716 | 0.19 | 0.178 | 1 |

**CD8+ T cells**

| | p_val | avg_logFC | IPF | Control | p_val_adj |
|---|---|---|---|---|---|
| MALAT1 | 0 | -0.3583805 | 1 | 0.999 | 0 |
| MT-ATP8 | 9.51E-204 | 0.26469429 | 0.972 | 0.927 | 2.23E-199 |
| RPS4Y1 | 2.47E-116 | 0.2528292 | 0.665 | 0.461 | 5.81E-112 |
| TRBV15 | 7.93E-105 | -0.4412252 | 0.007 | 0.052 | 1.86E-100 |
| HIST1H1E | 1.66E-86 | -0.4139879 | 0.208 | 0.317 | 3.90E-82 |
| MTRNR2L12 | 3.75E-75 | -0.2767186 | 0.909 | 0.925 | 8.81E-71 |
| TYROBP | 1.98E-62 | 0.37132997 | 0.231 | 0.139 | 4.65E-58 |
| HLA-DQB1 | 6.07E-56 | -0.2914706 | 0.133 | 0.207 | 1.42E-51 |
| TRBV3-1 | 2.44E-52 | 0.46054753 | 0.063 | 0.017 | 5.72E-48 |
| TRBV29-1 | 5.03E-35 | -0.3809718 | 0.024 | 0.055 | 1.18E-30 |
| S100B | 3.38E-27 | 0.34478049 | 0.053 | 0.023 | 7.92E-23 |
| TRBV19 | 1.34E-21 | 0.302249 | 0.058 | 0.029 | 3.15E-17 |
| TRBV20-1 | 4.03E-16 | -0.3591281 | 0.072 | 0.102 | 9.47E-12 |
| TRBV28 | 1.34E-15 | -0.3113056 | 0.046 | 0.071 | 3.14E-11 |
| DUSP2 | 8.64E-14 | -0.2724734 | 0.578 | 0.595 | 2.03E-09 |
| TRBV7-9 | 1.44E-10 | -0.3642423 | 0.043 | 0.062 | 3.38E-06 |
| MTRNR2L8 | 0.11164796 | 0.63048823 | 0.169 | 0.161 | 1 |

**Dendritic cells**

| | p_val | avg_logFC | IPF | Control | p_val_adj |
|---|---|---|---|---|---|
| ZFP36 | 9.01E-33 | -0.2593112 | 0.951 | 0.962 | 2.11E-28 |
| RPS4Y1 | 1.35E-30 | 0.28838156 | 0.74 | 0.463 | 3.17E-26 |
| HLA-DRB1 | 1.60E-28 | -0.2703714 | 0.996 | 0.992 | 3.76E-24 |
| MTRNR2L12 | 8.51E-21 | -0.3019794 | 0.97 | 0.982 | 2.00E-16 |
| DUSP2 | 4.18E-17 | -0.292248 | 0.397 | 0.522 | 9.80E-13 |
| SGK1 | 9.29E-17 | -0.3041042 | 0.346 | 0.453 | 2.18E-12 |
| GPR183 | 2.81E-14 | -0.2725214 | 0.48 | 0.581 | 6.58E-10 |
| BTG2 | 1.14E-13 | -0.251465 | 0.555 | 0.64 | 2.68E-09 |
| EGR1 | 1.59E-11 | -0.2665486 | 0.194 | 0.282 | 3.72E-07 |
| S100A12 | 1.08E-10 | 0.38635216 | 0.533 | 0.44 | 2.53E-06 |
| MT2A | 1.66E-10 | 0.30189516 | 0.602 | 0.521 | 3.90E-06 |
| S100A8 | 6.98E-09 | 0.32947463 | 0.729 | 0.655 | 0.00016385 |
| VCAN | 2.14E-07 | 0.25442211 | 0.617 | 0.574 | 0.00502078 |
| MTRNR2L8 | 0.76209151 | 0.57077451 | 0.317 | 0.324 | 1 |

**B cells**

| | p_val | avg_logFC | IPF | Control | p_val_adj |
|---|---|---|---|---|---|
| H3F3B | 0 | -0.518264 | 0.966 | 0.984 | 0 |
| EIF1 | 9.48E-216 | -0.2971937 | 0.993 | 0.994 | 2.22E-211 |
| MT-ATP8 | 1.32E-145 | 0.4204434 | 0.968 | 0.949 | 3.09E-141 |
| JUN | 2.99E-142 | -0.5306822 | 0.596 | 0.759 | 7.01E-138 |
| FOS | 9.02E-125 | -0.8635193 | 0.227 | 0.416 | 2.12E-120 |
| BTG2 | 1.29E-117 | -0.5130653 | 0.492 | 0.654 | 3.02E-113 |
| CD83 | 2.78E-117 | -0.525507 | 0.323 | 0.518 | 6.53E-113 |
| CD69 | 2.66E-112 | -0.7129086 | 0.385 | 0.561 | 6.24E-108 |
| KLF6 | 1.25E-108 | -0.435144 | 0.519 | 0.678 | 2.93E-104 |
| RGS2 | 4.33E-105 | -0.5507411 | 0.116 | 0.27 | 1.02E-100 |
| TENT5C | 8.16E-96 | -0.3953077 | 0.177 | 0.342 | 1.91E-91 |
| NR4A2 | 4.63E-92 | -0.474234 | 0.136 | 0.286 | 1.09E-87 |
| NR4A1 | 8.80E-92 | -0.4400968 | 0.033 | 0.134 | 2.06E-87 |
| JUNB | 9.23E-87 | -0.646413 | 0.854 | 0.851 | 2.16E-82 |
| AREG | 8.77E-79 | -0.4037061 | 0.069 | 0.183 | 2.06E-74 |
| RHOB | 9.17E-78 | -0.5625816 | 0.194 | 0.335 | 2.15E-73 |
| SBDS | 2.48E-70 | -0.3223011 | 0.251 | 0.396 | 5.81E-66 |
| DUSP1 | 1.30E-67 | -0.3959572 | 0.652 | 0.749 | 3.06E-63 |
| ZFP36 | 4.63E-65 | -0.4152451 | 0.738 | 0.779 | 1.09E-60 |
| MTRNR2L12 | 5.51E-63 | -0.2592724 | 0.931 | 0.969 | 1.29E-58 |
| ZNF331 | 1.08E-58 | -0.3148968 | 0.088 | 0.19 | 2.53E-54 |
| TSC22D3 | 7.94E-58 | -0.2628471 | 0.841 | 0.888 | 1.86E-53 |
| PELI1 | 1.75E-57 | -0.3156223 | 0.198 | 0.324 | 4.12E-53 |
| CXCR4 | 6.41E-54 | -0.338764 | 0.724 | 0.806 | 1.50E-49 |
| YPEL5 | 2.05E-48 | -0.2940574 | 0.311 | 0.422 | 4.81E-44 |
| IER2 | 2.97E-48 | -0.4698582 | 0.548 | 0.627 | 6.97E-44 |
| DUSP2 | 2.50E-47 | -0.386149 | 0.115 | 0.214 | 5.86E-43 |
| PPP1R15A | 1.29E-41 | -0.2836458 | 0.307 | 0.418 | 3.03E-37 |
| PDE4B | 2.46E-41 | -0.3044961 | 0.358 | 0.463 | 5.77E-37 |
| RHOH | 8.60E-36 | -0.2549209 | 0.489 | 0.577 | 2.02E-31 |
| HERPUD1 | 1.48E-33 | -0.313753 | 0.518 | 0.597 | 3.47E-29 |
| HIST1H1E | 7.21E-30 | -0.3309583 | 0.207 | 0.294 | 1.69E-25 |
| EGR1 | 3.87E-29 | -0.2635348 | 0.036 | 0.085 | 9.08E-25 |
| IFITM3 | 3.12E-27 | -0.2805192 | 0.204 | 0.283 | 7.33E-23 |
| SAT1 | 1.16E-25 | -0.2577091 | 0.546 | 0.608 | 2.71E-21 |
| GPR183 | 1.92E-25 | -0.2534129 | 0.182 | 0.257 | 4.50E-21 |
| IGLC2 | 1.38E-21 | -0.2793354 | 0.199 | 0.265 | 3.24E-17 |
| IGLC3 | 4.06E-08 | -0.3518129 | 0.211 | 0.25 | 0.0009522 |
| IGHV1-2 | 0.00011052 | -0.4106821 | 0.04 | 0.055 | 1 |
| IGLV1-51 | 0.00125849 | 0.5156281 | 0.051 | 0.037 | 1 |
| IGHV4-39 | 0.28077289 | 0.26636783 | 0.063 | 0.068 | 1 |
| IGKV1-5 | 0.33325615 | 0.4284374 | 0.076 | 0.071 | 1 |
| IGLV1-44 | 0.50001519 | -0.9633608 | 0.058 | 0.061 | 1 |

| | | | | | |
|---|---|---|---|---|---|
| IGHV3-21 | 0.56984385 | -0.293285 | 0.07 | 0.067 | 1 |
| IGHV3-33 | 0.99053363 | -0.2702246 | 0.052 | 0.052 | 1 |

**NK cells**

| | p_val | avg_logFC | IPF | Control | p_val_adj |
|---|---|---|---|---|---|
| MT-ATP8 | 3.91E-250 | 0.29668327 | 0.965 | 0.911 | 9.17E-246 |
| IL32 | 3.14E-157 | -0.4071408 | 0.395 | 0.575 | 7.36E-153 |
| KIR3DL1 | 8.67E-157 | -0.4199444 | 0.189 | 0.345 | 2.03E-152 |
| NFKBIA | 1.12E-131 | -0.3008819 | 0.54 | 0.668 | 2.63E-127 |
| KLRC1 | 2.06E-104 | 0.49378454 | 0.349 | 0.223 | 4.83E-100 |
| KIR2DL1 | 1.06E-103 | -0.322571 | 0.1 | 0.202 | 2.48E-99 |
| CMC1 | 6.14E-102 | 0.28674086 | 0.626 | 0.486 | 1.44E-97 |
| MYOM2 | 8.21E-99 | -0.6795189 | 0.183 | 0.291 | 1.93E-94 |
| BTG2 | 6.59E-93 | -0.2843443 | 0.208 | 0.328 | 1.55E-88 |
| CHI3L2 | 7.33E-91 | -0.2984262 | 0.061 | 0.139 | 1.72E-86 |
| HLA-DRB1 | 2.90E-83 | 0.38379041 | 0.278 | 0.167 | 6.79E-79 |
| HLA-DRB5 | 2.31E-58 | 0.25051995 | 0.165 | 0.088 | 5.43E-54 |
| TCF25 | 3.86E-55 | -0.2756825 | 0.51 | 0.58 | 9.04E-51 |
| HIST1H1E | 7.14E-45 | -0.2989491 | 0.275 | 0.357 | 1.67E-40 |
| PTGDS | 1.57E-41 | -0.307434 | 0.131 | 0.196 | 3.69E-37 |
| MTRNR2L8 | 9.73E-05 | 0.96869055 | 0.168 | 0.152 | 1 |
