## Supplementary Table E3 for "Single-cell profiling reveals immune aberrations in progressive idiopathic pulmonary fibrosis"

Supplementary Table E3: Differentially expressed genes in stable vs progressive IPF

**Monocytes**

| | p_val | avg_logFC | Progressive | Stable | p_val_adj |
|---|---|---|---|---|---|
| HLA-DQA2 | 0 | -0.4208707 | 0.099 | 0.258 | 0 |
| TMEM176B | 0 | -0.4215049 | 0.375 | 0.549 | 0 |
| CCL4 | 0 | -1.6165031 | 0.107 | 0.275 | 0 |
| TNFAIP3 | 3.34E-306 | -0.6947618 | 0.338 | 0.478 | 7.83E-302 |
| G0S2 | 6.94E-298 | -0.8308511 | 0.232 | 0.385 | 1.63E-293 |
| CCL4L2 | 2.67E-257 | -0.873981 | 0.063 | 0.173 | 6.27E-253 |
| CCL3 | 1.71E-207 | -0.9478196 | 0.252 | 0.379 | 4.01E-203 |
| TMEM176A | 3.38E-204 | -0.3033841 | 0.321 | 0.469 | 7.92E-200 |
| SAMSN1 | 4.00E-201 | -0.4033059 | 0.222 | 0.343 | 9.40E-197 |
| SOD2 | 5.95E-186 | -0.4578933 | 0.624 | 0.709 | 1.40E-181 |
| MTRNR2L8 | 3.32E-145 | 1.6974638 | 0.268 | 0.176 | 7.79E-141 |
| ICAM1 | 1.12E-141 | -0.2807127 | 0.243 | 0.349 | 2.63E-137 |
| IL1B | 6.78E-140 | -0.8507551 | 0.299 | 0.396 | 1.59E-135 |
| CCL3L1 | 1.14E-132 | -0.5157125 | 0.145 | 0.237 | 2.67E-128 |
| NFKBIA | 2.49E-130 | -0.5232407 | 0.869 | 0.882 | 5.83E-126 |
| NAMPT | 1.35E-129 | -0.2990371 | 0.708 | 0.771 | 3.17E-125 |
| CXCL2 | 1.91E-126 | -0.4948886 | 0.138 | 0.225 | 4.49E-122 |
| ACSL1 | 3.28E-125 | -0.3081972 | 0.276 | 0.375 | 7.70E-121 |
| RNF144B | 1.79E-121 | -0.3232758 | 0.336 | 0.428 | 4.20E-117 |
| LGALS2 | 7.76E-117 | 0.27473922 | 0.639 | 0.556 | 1.82E-112 |
| PLEK | 5.80E-114 | -0.3184186 | 0.756 | 0.798 | 1.36E-109 |
| CXCL3 | 2.93E-113 | -0.3241599 | 0.06 | 0.124 | 6.87E-109 |
| NLRP3 | 3.46E-113 | -0.2790666 | 0.289 | 0.382 | 8.11E-109 |
| CD83 | 1.72E-99 | -0.2832825 | 0.371 | 0.456 | 4.04E-95 |
| NFKBIZ | 4.31E-97 | -0.255158 | 0.689 | 0.738 | 1.01E-92 |
| BCL2A1 | 3.85E-92 | -0.3373849 | 0.528 | 0.598 | 9.04E-88 |
| CXCL8 | 4.87E-83 | -0.3210818 | 0.353 | 0.438 | 1.14E-78 |
| FOLR3 | 1.32E-69 | 0.53690904 | 0.314 | 0.258 | 3.10E-65 |
| TNF | 1.03E-66 | -0.3187922 | 0.206 | 0.271 | 2.41E-62 |
| JUN | 1.60E-56 | 0.39560714 | 0.505 | 0.45 | 3.74E-52 |
| THBS1 | 4.69E-51 | -0.2878129 | 0.196 | 0.259 | 1.10E-46 |
| GNLY | 7.10E-27 | -0.3506146 | 0.054 | 0.081 | 1.67E-22 |

**CD4+ T cells**

| | p_val | avg_logFC | Progressive | Stable | p_val_adj |
|---|---|---|---|---|---|
| HLA-B | 0 | 0.36979387 | 0.999 | 0.997 | 0 |
| LGALS1 | 2.29E-69 | 0.29678405 | 0.379 | 0.261 | 5.38E-65 |
| IFITM3 | 3.39E-68 | -0.2512314 | 0.287 | 0.392 | 7.95E-64 |
| MTRNR2L8 | 1.02E-27 | 1.36133944 | 0.215 | 0.163 | 2.39E-23 |
| MT2A | 5.13E-19 | 0.37262059 | 0.399 | 0.349 | 1.20E-14 |
| GADD45B | 3.84E-13 | 0.42384765 | 0.361 | 0.32 | 9.00E-09 |
| JUN | 0.03832919 | 0.27941129 | 0.314 | 0.306 | 1 |

**CD8+ T cells**

| | p_val | avg_logFC | Progressive | Stable | p_val_adj |
|---|---|---|---|---|---|
| RPS26 | 0 | -0.732769 | 0.954 | 0.992 | 0 |
| IFITM3 | 1.57E-181 | -0.5271247 | 0.288 | 0.517 | 3.69E-177 |
| GNLY | 6.17E-137 | -0.521988 | 0.505 | 0.686 | 1.45E-132 |
| FGFBP2 | 4.20E-102 | -0.3882577 | 0.475 | 0.636 | 9.86E-98 |
| IER2 | 1.06E-99 | -0.374624 | 0.674 | 0.767 | 2.49E-95 |
| NFKBIA | 3.72E-94 | -0.6435516 | 0.589 | 0.66 | 8.72E-90 |
| HOPX | 3.32E-90 | -0.2752729 | 0.508 | 0.68 | 7.80E-86 |
| CD27 | 8.41E-78 | 0.2935115 | 0.333 | 0.196 | 1.97E-73 |
| JUN | 1.05E-76 | -0.4823089 | 0.215 | 0.348 | 2.47E-72 |
| ZNF683 | 2.41E-69 | -0.3959063 | 0.134 | 0.257 | 5.65E-65 |
| TNFAIP3 | 3.48E-69 | -0.3883176 | 0.268 | 0.397 | 8.16E-65 |
| ID2 | 6.03E-58 | -0.2649072 | 0.418 | 0.544 | 1.42E-53 |
| S100A8 | 1.94E-49 | 0.30031013 | 0.247 | 0.152 | 4.55E-45 |
| TRBV7-9 | 8.58E-47 | 0.35083179 | 0.075 | 0.024 | 2.01E-42 |
| HLA-DRB5 | 3.55E-46 | -0.2851265 | 0.232 | 0.338 | 8.33E-42 |
| GZMK | 1.31E-41 | 0.29118619 | 0.44 | 0.332 | 3.07E-37 |
| TRBV19 | 8.79E-41 | 0.42189957 | 0.091 | 0.038 | 2.06E-36 |
| TRBV4-2 | 7.18E-33 | -0.4687065 | 0.013 | 0.052 | 1.69E-28 |
| ZFP36 | 2.32E-31 | -0.2574325 | 0.653 | 0.701 | 5.45E-27 |
| TRBV7-6 | 6.84E-25 | 0.3913097 | 0.078 | 0.039 | 1.61E-20 |
| LTB | 1.11E-24 | 0.28581091 | 0.439 | 0.366 | 2.59E-20 |
| MT2A | 5.49E-23 | 0.28132825 | 0.585 | 0.513 | 1.29E-18 |
| TRBV28 | 2.99E-19 | -0.2718973 | 0.026 | 0.058 | 7.01E-15 |
| CXCR4 | 7.45E-15 | -0.2776073 | 0.493 | 0.518 | 1.75E-10 |
| MTRNR2L8 | 1.30E-14 | 1.3355832 | 0.194 | 0.154 | 3.05E-10 |
| TRBV5-1 | 4.41E-14 | 0.32882739 | 0.052 | 0.028 | 1.04E-09 |
| TRBV2 | 1.62E-10 | -0.3644167 | 0.031 | 0.054 | 3.79E-06 |

**Dendritic cells**

| | p_val | avg_logFC | Progressive | Stable | p_val_adj |
|---|---|---|---|---|---|
| HLA-DQA2 | 3.81E-28 | -0.767906 | 0.371 | 0.521 | 8.94E-24 |
| CCL4 | 3.30E-06 | -0.4058287 | 0.049 | 0.098 | 0.07745646 |
| CCL3 | 7.09E-06 | -0.399869 | 0.103 | 0.159 | 0.16636421 |
| IL1B | 1.24E-05 | -0.3172354 | 0.307 | 0.376 | 0.29138503 |
| AREG | 0.08037412 | -0.3021383 | 0.244 | 0.274 | 1 |
| MTRNR2L8 | 0.53980007 | 0.99281082 | 0.318 | 0.315 | 1 |

**B cells**

| | p_val | avg_logFC | Progressive | Stable | p_val_adj |
|---|---|---|---|---|---|
| RPS26 | 2.14E-285 | -0.7080274 | 0.868 | 0.977 | 5.01E-281 |
| HLA-C | 3.49E-125 | -0.3888749 | 0.947 | 0.985 | 8.20E-121 |
| IGHG2 | 7.24E-30 | -0.2994645 | 0.156 | 0.272 | 1.70E-25 |
| MARCKSL1 | 2.41E-29 | 0.29619828 | 0.425 | 0.304 | 5.65E-25 |
| HERPUD1 | 2.65E-28 | -0.3222289 | 0.46 | 0.561 | 6.22E-24 |
| JUNB | 2.94E-20 | -0.2680158 | 0.834 | 0.869 | 6.90E-16 |
| CD83 | 1.40E-16 | -0.3562801 | 0.277 | 0.357 | 3.27E-12 |
| IGHA1 | 7.10E-10 | -0.5854431 | 0.165 | 0.225 | 1.67E-05 |
| IGLV3-1 | 7.17E-08 | 0.29132137 | 0.081 | 0.048 | 0.00168274 |
| IGHV4-34 | 3.38E-07 | 0.69381851 | 0.077 | 0.048 | 0.00792001 |
| IGHG1 | 4.58E-07 | -0.3094694 | 0.102 | 0.144 | 0.01073799 |
| IGHV3-7 | 6.95E-05 | -0.6353777 | 0.035 | 0.056 | 1 |
| IGKV3-20 | 0.0001129 | 0.7854279 | 0.143 | 0.111 | 1 |
| IGKV4-1 | 0.00018457 | 0.71964 | 0.128 | 0.097 | 1 |
| IGLV1-51 | 0.00118265 | -0.6005662 | 0.041 | 0.058 | 1 |
| IGHV4-39 | 0.00998312 | 0.86223405 | 0.072 | 0.057 | 1 |
| IGLV3-21 | 0.01459838 | -0.5465481 | 0.041 | 0.054 | 1 |
| IGKV3-11 | 0.09275463 | 0.47945697 | 0.083 | 0.072 | 1 |
| IGLC2 | 0.30935016 | -0.3982792 | 0.194 | 0.203 | 1 |
| IGHV4-31 | 0.31926181 | 0.33695909 | 0.051 | 0.046 | 1 |
| IGHV5-51 | 0.5094457 | 0.2789678 | 0.064 | 0.06 | 1 |
| IGHV3-21 | 0.64441495 | -0.2695697 | 0.069 | 0.071 | 1 |
| IGLV1-44 | 0.68919516 | -0.547597 | 0.06 | 0.057 | 1 |

**NK cells**

| | p_val | avg_logFC | Progressive | Stable | p_val_adj |
|---|---|---|---|---|---|
| HLA-B | 0 | 0.30145095 | 1 | 1 | 0 |
| IGFBP7 | 1.33E-129 | 0.34077654 | 0.677 | 0.495 | 3.12E-125 |
| FCGR3A | 1.36E-105 | 0.27841945 | 0.894 | 0.853 | 3.18E-101 |
| MTRNR2L8 | 4.10E-97 | 1.85784111 | 0.241 | 0.127 | 9.62E-93 |
| MYOM2 | 4.21E-95 | 0.46644468 | 0.265 | 0.138 | 9.89E-91 |
| HLA-DRB1 | 1.20E-86 | -0.4922242 | 0.194 | 0.324 | 2.81E-82 |
| CD52 | 3.94E-77 | -0.285305 | 0.619 | 0.725 | 9.25E-73 |
| TRAC | 2.79E-73 | -0.3480339 | 0.055 | 0.146 | 6.54E-69 |
| HLA-DRB5 | 2.26E-70 | -0.3565999 | 0.099 | 0.202 | 5.31E-66 |
| HLA-DPA1 | 5.39E-67 | -0.3241231 | 0.367 | 0.479 | 1.26E-62 |
| HLA-DPB1 | 8.67E-62 | -0.2953559 | 0.389 | 0.499 | 2.03E-57 |
| S100A9 | 1.60E-49 | 0.29180189 | 0.29 | 0.192 | 3.76E-45 |
| NFKBIA | 2.47E-45 | -0.4352494 | 0.497 | 0.563 | 5.81E-41 |
| S100B | 1.02E-38 | 0.38583069 | 0.134 | 0.072 | 2.40E-34 |
| IL32 | 4.52E-30 | -0.2549207 | 0.339 | 0.425 | 1.06E-25 |
| SELL | 1.52E-29 | 0.2869115 | 0.305 | 0.23 | 3.57E-25 |
| KLRC1 | 3.49E-29 | -0.3379641 | 0.308 | 0.372 | 8.19E-25 |
| MT2A | 7.46E-27 | 0.31768365 | 0.565 | 0.494 | 1.75E-22 |
| GZMK | 1.31E-19 | -0.2622395 | 0.061 | 0.101 | 3.06E-15 |
| CD74 | 1.29E-15 | -0.2729808 | 0.86 | 0.852 | 3.03E-11 |
| TKTL1 | 1.23E-09 | -0.2521767 | 0.06 | 0.085 | 2.89E-05 |
| JUN | 0.00109291 | -0.260397 | 0.133 | 0.146 | 1 |
