## Supplementary Table E4 for "Single-cell profiling reveals immune aberrations in progressive idiopathic pulmonary fibrosis"

Supplementary Table E4: List of antibodies used in CyTOF

| | Metal | Target | Catalog # | Clone | Vendor ID |
|---|---|---|---|---|---|
| 1 | 89Y | CD45 | 3089003B | HI30 | Fluidigm/DVS |
| 2 | 141Pr | CD235ab | 3141001B | HIR2 | Fluidigm/DVS |
| 3 | 142Nd | CD19 | 3142001B | HIB19 | Fluidigm/DVS |
| 4 | 146Nd | CD8a | 3146001B | RPA-T8 | Fluidigm/DVS |
| 5 | 147Sm | CD20 | 3147001B | 2H7 | Fluidigm/DVS |
| 6 | 148Nd | CD16 | 3148004B | 3G8 | Fluidigm/DVS |
| 7 | 149Sm | CD66 | 3149008B | CD66a-B1.1 | Fluidigm/DVS |
| 8 | 150Nd | CD194(CCR4) | V04603 | L291H4 | LW |
| 9 | 151Eu | CD123 | 3151001B | 6H6 | Fluidigm/DVS |
| 10 | 153Eu | IgD | V05298 | IA6-2 | LW |
| 11 | 155Gd | CD4 | V04286 | RPA T4 | LW |
| 12 | 156Gd | CD183(CXCR3) | 3156004B | CXCR3(G025H7) | Fluidigm/DVS |
| 13 | 159Tb | CD11c | 3159001B | Bu15 | Fluidigm/DVS |
| 14 | 160Gd | CD14 | 3160001B | M5E2 | Fluidigm/DVS |
| 15 | 164Dy | CD45RO | 3164007B | UCHL1 | Fluidigm/DVS |
| 16 | 165Ho | Foxp3 | V03318 | PCH101 | LW |
| 17 | 167Er | CD27 | 3167002B | O323 | Fluidigm/DVS |
| 18 | 168Er | CD196(CCR6) | V05986 | G034E3 | LW |
| 19 | 169Tm | CD25 | 3169003B | 2A3 | Fluidigm/DVS |
| 20 | 171Yb | CD127 | | A019D5 | LW |
| 21 | 172Yb | CD38 | 3172007B | HIT2 | Fluidigm/DVS |
| 22 | 174Yb | HLA-DR | 3174001B | L243 | Fluidigm/DVS |
| 23 | 176Yb | CD56 | 3176009B | N901 | Fluidigm/DVS |
| 24 | 158Gd | CD3 | | UCHT1 | LW |
| 25 | 143Nd | CD45RA | 3143006B | Hi100 | Fluidigm/DVS |
| 26 | 162Dy | pLck [T505] | 3162004A | 4/LCK-Y505 | Fluidigm/DVS |
| 27 | 209Bi | CD61 | 3209001B | VI-PL2 | Fluidigm/DVS |
| 28 | 163Dy | TGFbeta | 3163010B | TW4-6H10 | Fluidigm/DVS |
| 29 | 170Er | CCR7(CD197) | | G043H7 | LW |
